# Exome sequencing in Asian populations identifies rare deficient *SMPD1* alleles that increase risk of Parkinson’s disease

**DOI:** 10.1101/2023.08.03.23293387

**Authors:** Elaine GY Chew, Zhehao Liu, Zheng Li, Sun Ju Chung, Michelle M Lian, Moses Tandiono, Ebonne Y Ng, Louis CS Tan, Wee Ling Chng, Tiak Ju Tan, Esther KL Peh, Ying Swan Ho, Xiao Yin Chen, Erin YT Lim, Chu Hua Chang, Jonavan J Leong, Yue Jing Heng, Ting Xuan Peh, Ling-Ling Chan, Yinxia Chao, Wing-Lok Au, Kumar M Prakash, Jia Lun Lim, Yi Wen Tay, Vincent Mok, Anne YY Chan, Juei-Jueng Lin, Beom S Jeon, Kyuyoung Song, Clement CY Tham, Chi Pui Pang, Jeeyun Ahn, Kyu Hyung Park, Janey L Wiggs, Tin Aung, Ai-Huey Tan, Azlina Ahmad Annuar, Mary B. Makarious, Cornelis Blauwendraat, Mike A Nalls, Laurie A. Robak, Roy N. Alcalay, Ziv Gan-Or, Shen-Yang Lim, Chiea Chuen Khor, Eng-King Tan, Zhenxun Wang, Jia Nee Foo

## Abstract

Parkinson’s disease is an incurable and progressive disease that adversely affects balance, muscle control, and movement. We hypothesized that the landscape of rare, protein-altering genetic variants could provide further mechanistic insights into disease pathogenesis. We performed whole-exome sequencing on 4,298 persons with Parkinson’s disease and 5,512 unaffected controls from Singapore, Malaysia, Hong Kong, South Korea, and Taiwan. We tested for association between gene-based burden of rare, predicted damaging variants and risk of Parkinson’s disease. Genes surpassing exome-wide significance (*P*<2.5×10^-6^) were tested for replication in sequencing data from a further 5,585 Parkinson’s disease patients and 5,642 controls of Asian and European ancestry. We observed that carriage of rare, protein-altering variants that were predicted to impair protein function at *SMPD1* (a gene encoding for acid sphingomyelinase) were significantly associated with increased risk of Parkinson’s disease. Refinement of variant classification using functional acid sphingomyelinase assays suggest that individuals carrying *SMPD1* variants with less than 44 percent of normal enzymatic activity show the strongest association with Parkinson’s disease risk in both the discovery (odds ratio (OR) = 2.37, 95% CI = 1.68 - 3.35, *P* = 4.35 × 10^-7^) and replication collections (OR = 2.18, 95% CI = 1.69 - 2.81, *P* = 4.80 × 10^-10^), leading to a significant observation when all data were meta-analyzed (OR = 2.24, 95% CI = 1.83 - 2.76, *P* = 1.25 × 10^-15^). Our findings affirm the importance of sphingomyelin metabolism in the pathobiology of neurodegenerative diseases and highlights the utility of functional genomic assays in large-scale exome sequencing studies.

## Introduction

Parkinson’s disease is a common neurodegenerative disease that is unrelenting and progressive. It presents as a movement disorder characterized by slowness of movement, tremors, stiffness, and difficulty with balance, together with a broad array of non-motor symptoms^1,2^. Although levodopa and other adjunctive medications alleviate the symptoms of Parkinson’s disease, no cure has yet been found. There is thus a pressing need for new therapeutic approaches, with the hope that biological insights arising from molecular genetic studies could illuminate these otherwise elusive drug targets.

Parkinson’s disease has long been suspected to be heritable due to reports of positive family histories in many affected individuals. To this end, genome-wide association studies (GWAS) of Parkinson’s disease have identified close to 100 loci associated with disease risk^3-9^. While these studies highlighted key genes and biological pathways that play a role in the pathogenesis of Parkinson’s disease, they also underscore the very heterogeneous nature of the disease in terms of causative mechanisms.

The genomic/risk loci identified by GWAS of Parkinson’s disease mostly have modest effect sizes (odds ratios of less than 1.4) and were often located in non-coding regions of the human genome^9^. Their non-coding nature makes pinpointing the effector genes challenging. In contrast, analysis of protein-altering genetic variants using exome sequencing has identified causative genes and facilitated advances in the treatment of other diseases through the identification and development of new drug targets^10,11^. In studies on familial, early-onset Parkinson’s disease, several genes containing rare functionally deficient genetic variants have been found^12-26^. Such variants were highly penetrant and were seldom seen in patients with sporadic Parkinson’s disease^27^. In the case of adult-onset sporadic Parkinson’s disease, the effect of individual rare genetic variants remains difficult to assess, but the aggregation of rare variant counts within each gene could allow for detection of statistically significant differences in mutational ‘burden’ between cases and controls. However, gene-based tests of mutational burden are often limited by constraints in accuracy of pathogenicity prediction using bioinformatic algorithms^28^ because not all variants that are predicted to impair protein function actually do so. Conversely, variants that are predicted to be benign could result in a loss of function of the encoded protein.

The objective of this study was to assess the contribution of rare coding-sequence genetic variants to the pathogenesis of Parkinson’s disease. We performed whole-exome sequencing on 9,810 participants of Asian ancestry (4,298 persons with Parkinson’s disease and 5,512 unaffected controls) from Singapore, Malaysia, Hong Kong, South Korea, and Taiwan. We attempted to replicate significantly associated genes in a further 11,227 participants (5,585 Parkinson’s disease patients and 5,642 controls) from three independently ascertained collections of Asian and European ancestry. We used functional biological assays to improve upon gene-based mutational burden tests.

## Methods

### Study design and participants

#### Discovery exome sequencing

Patients with Parkinson’s disease together with ancestry- and geographically matched controls were recruited by 6 independent study groups from 5 regions across East Asia (Singapore (SG), Malaysia (MAL), Hong Kong (HK), South Korea (KR) and Taiwan (TW)). A total of 4,298 cases and 5,512 controls were included in the final analysis (**Table S1**, **Table S2A**). A diagnosis of Parkinson’s disease was reached using the United Kingdom Parkinson’s Society Brain Bank Criteria. Conversely, the controls were healthy subjects that do not have any overt neurological diseases. Informed consent was obtained from each participant in strict adherence to the Declaration of Helsinki. This study was approved by the ethics committees or institutional review boards of the respective institutions (SingHealth Centralized Institutional Review Board CIRB 2002/008/A and 2019/2334 and Nanyang Technological University Institutional Review Board IRB-2016-08-011). The study was conducted from January 1, 2016, to December 31, 2022. This study followed the Strengthening the Reporting of Genetic Association Studies (STREGA) reporting guideline.

#### Replication datasets and analysis

Published sequencing data from a total of 5,585 Parkinson’s disease patients and 5,642 controls from China, United States, and the International Parkinson’s Disease Genomics Consortium (samples ascertained from the United States and from Europe) were evaluated for replication of significant findings from the discovery exome sequencing analysis (**Table S2B**).

For the Chinese dataset, a total of 1,440 persons with sporadic early-onset Parkinson’s disease, 477 persons with a positive family history of Parkinson’s disease, 1,962 persons with sporadic, late-onset Parkinson’s disease, and 2,931 unaffected control individuals from Changsha were included^29^ (referred to as the “Zhao” dataset/stratum). For the International Parkinson’s Disease Genomics Consortium dataset, a total of 691 persons with sporadic early onset Parkinson’s disease, 465 persons with a positive family history of Parkinson’s disease, and 1,679 unaffected control individuals of Northern and Western European ancestry were included^30^ (referred to as the “Robak” dataset/stratum). For the United States dataset, a total of 550 patients with Parkinson’s disease and 284 controls from New York were enrolled^31^. This dataset was enriched with an additional 748 unaffected control individuals from neighboring Boston, USA ^32^ (referred to as the “Alcalay-Boston” dataset/stratum). Carrier counts of rare damaging *SMPD1* variants among cases and controls were meta-analyzed for each of these three replication collections using the stratified Cochran-Mantel-Haenszel (CMH) test without continuity correction.

### Whole-exome capture and sequencing

Peripheral venous blood was drawn from each consenting participant and DNA extraction was performed using routine laboratory methods. DNA for whole-exome sequencing was sheared using Covaris LE220 Focused-ultrasonicator and libraries were prepared using NEBNext Ultra II DNA Library Prep Kit for Illumina (New England Biolabs, USA). Exomes were captured using SeqCap EZ Human Exome Library v3.0 kits or KAPA HyperExome Probes (both from Roche NimbleGen Inc., USA). Libraries from individuals with Parkinson’s disease and unaffected individuals were barcoded and sequenced together in batches of 96 samples using 2 × 151 bp paired-end reads on Illumina instruments (HiSeq 4000 and NovaSeq 6000 systems) to achieve at least 60x average depth across captured target regions. All samples were processed in the same laboratory.

### Bioinformatics processing, variant calling and quality control filtering

For the discovery exome sequencing dataset, raw DNA sequence reads from all samples (with case-control status blinded) were mapped to the Human reference genome build Hg19 (GRCh37) using the Burrows-Wheeler Aligner MEM algorithm (BWA version 0.7.17) (**Figure S1**). Variant calling was performed using the Genome Analysis Tool Kit (GATK, version 3.7)^33^ and the Picard (version 2.21.7) (http://broadinstitute.github.io/picard/) software packages (Broad Institute) following GATK Best Practices Guidelines^34^. Variant quality score recalibration (VQSR) was performed to exclude low quality SNP calls.

Additional quality control procedures were performed using VCFtools (version 0.1.16)^35^, BCFtools (version 1.9)^36^, PLINK (version 1.9)^37^ and the Fratools PERL package (https://github.com/atks/fratools) at genotype, variant call and sample quality check levels. Low quality genotype calls (DP <8 and GQ <20) were excluded. Variants with low call rates in cases, controls and all samples respectively (<95%), failed tests for Hardy-Weinberg equilibrium (*P*<1×10^-6^) and high differential missingness between cases and controls (*P*<1×10^-4^) were excluded from further analyses. We removed patients with sex discordance between genetically inferred sex obtained from exome sequencing with patient records. We also removed samples showing poor genotyping concordance (<95% concordance) between exome sequence data and previous fingerprinting with genome-wide association arrays. Samples with excessive heterozygosity (defined as >3.5 standard deviations from the mean), unusually high exome-wide singleton counts, and low genotyping completion rate (<95%) were also excluded. We used identity-by-descent analysis to identify related sample pairs up to the third degree (IBD >0.125). For these pairs, the sample with the lower call rate was removed. Lastly, samples with outlying genetic ancestry as visualized using ancestry principal component analysis were also removed. We took care to treat variant identification in all samples equally regardless of affection status. All steps related to sample quality checks are summarized in **Table S3**.

### Identification of Rare Damaging Genetic Variants

Single nucleotide variants and small insertions and deletions were annotated using the Ensembl Variant Effect Predictor (VEP) software (version 104)^38^. Variant annotation was based on canonical transcripts as defined in Ensembl VEP. Rare variants were defined based on allele frequency annotations (≤1%) from the exome sequencing dataset as well as from the publicly available Genome Aggregation Database (gnomAD) East Asian population^39^. Functional predictions for missense variants were derived from PolyPhen-2 (version 2.2.3r406)^40^ while the VEP plugin LOFTEE (version 1.0.3)^41^ was used for functional prediction of stop gain or stop loss, splice site disrupting and frameshift variants. Each variant was classified as damaging based on PolyPhen2 (probably or possibly damaging) and LOFTEE (high confidence) predictions. For the replication datasets, published variant lists from whole-exome and whole-genome sequencing of *GBA1* and *SMPD1* were re-annotated based on the listed variant position in Hg19/GRCh37 and allele information in the same way as the discovery dataset using PolyPhen-2 and Ensembl VEP software.

### Validation by Sanger Capillary Sequencing

DNA samples from participants carrying rare *SMPD1* alleles were selected for PCR amplification and capillary sequencing to confirm the rare allele calls from whole exome sequencing. PCR primers were designed using Primer3 (https://primer3.ut.ee/). Primer sequences and PCR conditions are listed in **Table S4**.

### Statistical analysis

Rare damaging variants within each gene were aggregated together for an association analysis using the burden test as previously described^32^. We use the burden test for primary analysis because many medical conditions show a haploinsufficiency effect. Haploinsufficiency refers to a state where one copy of a gene has been damaged by mutations, leaving the remaining normally functioning copy insufficient in sustaining normal function. The burden test is modeled on the hypothesis that different mutations within a gene cause the same damaging consequences to the encoded protein product.

We tested whether any of the 17,396 autosomal genes across the human exome bear an excess or deficit of ‘damaging’ mutational burden in persons with Parkinson’s disease compared to unaffected individuals. For the discovery dataset, a stratified CMH test was used to evaluate gene-based burden across the exomes of the study participants from the 5 countries studied (**Table S2A**). Singapore and Malaysia samples were considered as one stratum due to the similarity in genetic background (as evidenced by PCA of Singapore and Malaysia samples, **Figure S2**). We also evaluated gene-based burden after adjustment of covariates such as principal components (PC1-PC3), per sample variant counts and average per sample coverage. Exome-wide significance was preset at P < 2.5 × 10^−6^ (two-tailed), taking into account multiple hypothesis testing correction for an estimated 20,000 protein-coding genes in the human genome. Correlation of *SMPD1* and *GBA1* variant carrier status with age of onset of Parkinson’s disease was conducted with t-test on 4,149 out of 4,298 patients with Parkinson’s disease that have available information on age of disease onset.

For the replication stage, the same CMH test was used to summarize gene-based burden test results for each of the datasets from China, USA, and the International Parkinson’s Disease Genomic Consortium in a stratified meta-analysis in the same manner as was performed for the discovery stage.

### Acid sphingomyelinase enzymatic activity assays

*SMPD1* encodes for enzyme acid sphingomyelinase (ASM) which breaks down sphingomyelin to ceramide and phosphorylcholine^42^ To assess ASM activity associated with *SMPD1* variants observed, we generated HEK 293 cell lines that contain individual *SMPD1* variants (Table S5A) and conducted a fluorescence-based assay to measure ASM activity. We transduced HEK 293 cell lines with individual *SMPD1* variants cloned into the lentiviral pLVX-CMV vector containing a puromycin selection marker (**Supplemental Methods**)^43^ by standard lentiviral transduction methods. Cells were first transduced and then subsequently grown in puromycin containing media for a further 4 days until selection was complete. Western blotting was conducted with anti-SMPD1 antibody (#MAB5348, R&D Systems) to verify successful transduction by ascertainment of SMPD1 expression. A total of 125 cell lines expressing 125 individual protein-altering *SMPD1* genetic variants were made (122 rare variants, 3 common variants, **Table S5B**).

To measure ASM enzymatic activity associated with each *SMPD1 genetic* variant, cells were first lysed by sonication in water with protease inhibitor (#05056489001, Roche). Resultant cell extracts were then quantified for protein amount (#23200, Thermofisher) and 10 μg of extract were tested for ASM activity in replicates (n=3-6), using a fluorescence assay utilizing the synthetic substrate HMU-PC (#EH31028, Biosynth)^44^. We measured the ASM enzymatic activity level of each *SMDP1* allele as the amount of fluorescent signal generated over 1 hour at 37℃. To correct for batch-to-batch variation, a reference cell line expressing wild-type *SMPD1* alleles was always assayed together with the cell lines bearing *SMPD1* variant alleles. This allowed for the fluorescent signal of the cell extracts bearing *SMPD1* variants to be directly compared to the fluorescent signal of the cell lysates from the reference cell line expressing wild-type *SMPD1* alleles within each batch of experiment. Measured activity for each variant was presented as a functional score that was calculated based on log2 fold change of signal per replicate by the average wild-type signal.

To assess passage-to-passage activity variation, we randomly selected 8 cell lines containing 8 different *SMPD1* variants, generated lysates from them, and then measured their resultant ASM enzymatic activities using the same assay. The measured ASM enzymatic activity of all 8 variants relative to wild-type *SMPD1* remained highly concordant between passages (**Figure S3**).

### Analysis of acid sphingomyelinase enzymatic activity in discovery and replication datasets

Enzymatic activity of 125 tested rare variants (45 from the discovery exome sequencing stage, and 100 from the replication stage, with 20 variants found in both Discovery and Replication) were compared with enzymatic activity of wild-type *SMPD1* and functional scores were assigned based on percentage in reduction of activity. We explored the correlation of functional scores in relation to amino acid position on ASM protein structure^45,46^. Because ASM enzymatic activity was measured as a continuous variable, we used iterative linear discriminant analysis to explore enzymatic activity thresholds in which variants below a tested threshold would be classified as ‘impaired function’. This was done in order to obtain the best discrimination (in terms of Odds Ratios) between participants with Parkinson’s disease and unaffected controls. Only functionally impaired variants with enzymatic activities below the defined threshold were included in the revised gene-based burden test for *SMPD1*. The revised burden test was the same stratified CMH test used in the primary analysis.

## Results

### Whole exome sequencing identified rare coding variants in GBA1 and SMPD1 associated with Parkinson’s disease

We analyzed a total of 4,298 PD cases and 5,512 controls by whole exome sequencing (total N = 9,810, **Table S2A**). We identified a total of 721,643 non-synonymous variants across 17,396 autosomal genes, of which 701,048 of these non-synonymous variants were rare (minor allele frequency <1%) in the sequenced dataset as well as in publicly available datasets (gnomAD East Asian samples). After annotation with the PolyPhen-2 and LOFTEE prediction algorithms, 367,878 potentially pathogenic variants (thereafter termed ‘qualifying variants’) remained for further analysis.

Gene-level association testing revealed exome-wide significant associations between carriage of qualifying variants in *GBA1* and *SMPD1* (**Figure 1**). At *GBA1*, an aggregate of 128 out of 4,298 participants with Parkinson’s disease (2.98%) carried qualifying variants compared to 16 out of 5,512 unaffected controls (0.29%; OR = 10.28, 95% CI = 5.96-17.75, *P* = 4.07 × 10^-25^ **Table S6A**, **Figure S4, Figure S5**). Turning to *SMPD1*, 100 out of 4,298 participants with Parkinson’s disease (2.33%) carried qualifying variants compared to 69 out of 5,512 unaffected controls (1.25%; OR = 2.32, 95% CI = 1.68-3.20, *P* = 1.37 × 10^-7^, **Table 1**, **Figure 1, Figure S6**). In contrast, we did not observe significant enrichment of *GBA1* and *SMPD1* variants that were predicted to be benign in participants with Parkinson’s disease compared to controls for GBA1 (*P* = 0.05, OR = 1.36, **Table S6A**) or *SMPD1* (*P* = 0.70, OR = 1.11, **Table 1**). We also did not observe significant enrichment of rare synonymous variants in *GBA1* (*P* = 0.30, OR = 1.73, **Table S6A**) or *SMPD1* (*P* = 0.38, OR = 1.25, **Table S6B**) in patients with Parkinson’s disease compared to controls. Both *GBA1* and *SMPD1* were sequenced to > 60x coverage, with > 99% of targeted exons covered at least 10 times. These findings suggest that the exome-wide significant association between both genes and Parkinson’s disease were unlikely to be due to sequencing artifacts such as differential sequencing depth between participants with Parkinson’s disease and controls. We did not observe significant association at genes previously implicated in autosomal dominant, recessive, or X-linked Parkinson’s disease (**Table S7A**)^47-49^ or novel genes identified by a recent whole exome sequencing study in participants of European ancestry (**Table S7B**)^27^. We also did not observe significant associations (*P*< 0.00058; Bonferroni correction for 86 tested genes) of qualifying variants in lysosomal storage disorders in persons with Parkinson’s disease compared to controls (**Table S7C**)^29,30^.

**Figure 1:**
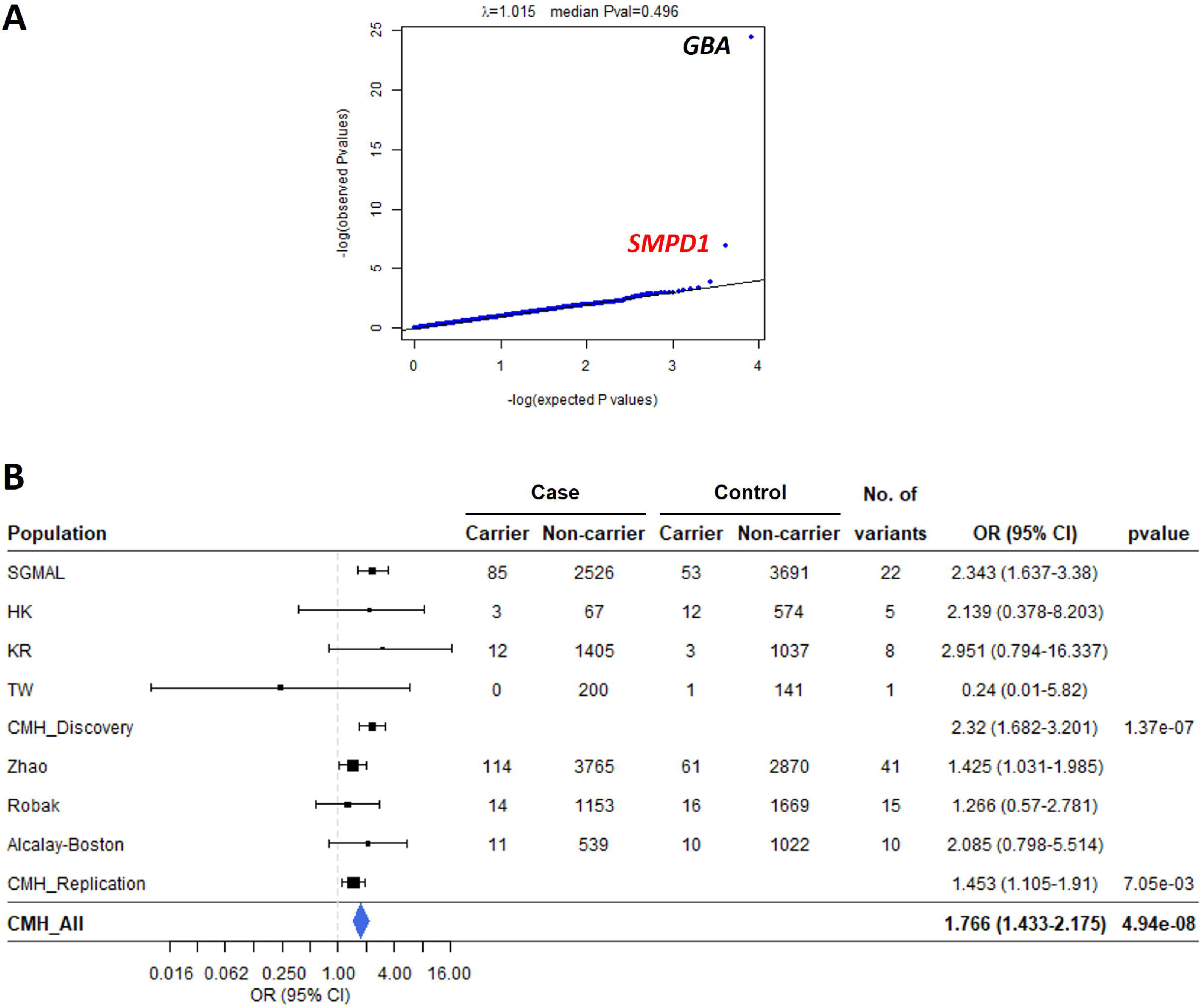
Exome-wide significant association of *SMPD1* in the discovery exome-sequencing study. (A) Quantile-Quantile plot of P-values for autosomal genes with OR > 1. (B) Forest plot of *SMPD1* in assessed populations. Error bars represent 95% confidence interval (CI).

**Table 1:** *SMPD1* coding variants from the Discovery dataset.

|  |  | gnomAD2.1.1 EAS frequencies |  | Case |  | Control |  |  |  |
| --- | --- | --- | --- | --- | --- | --- | --- | --- | --- |
| Pos | AA Substitution | exomes | genomes | no. of heterozygotes | no. of homozygotes | no. of heterozygotes | no. of homozygotes | Pathogenicity prediction | Functional score |
| Rare deleterious variants |  |  |  |  |  |  |  |  |  |
| chr11.6411889.C.T | p.Gln21Ter | NA | NA | 0 | 0 | 1 | 0 | Stopgain | 25.00% (24.90% - 25.10%) |
| chr11.6412063.C.T | p.Arg79Trp | NA | NA | 1 | 0 | 0 | 0 | Probably damaging | 69.60% (69.30% - 69.80%) |
| chr11.6413101.C.T | p.Ala269Val | 4.00E-04 | 6.00E-04 | 1 | 0 | 2 | 0 | Probably damaging | 98.80% (92.30% - 105.70%) |
| chr11.6413106.C.A | p.Pro271Thr | NA | NA | 0 | 0 | 1 | 0 | Damaging | 76.40% (75.30% - 77.50%) |
| chr11.6413134.A.ACATCCCCG | p.His284SerfsTer18 | NA | NA | 2 | 0 | 0 | 0 | Frameshift insertion | 5.60% (5.40% - 5.90%) |
| chr11.6413137.TC.T | p.Ala283HisfsTer16 | NA | NA | 0 | 0 | 1 | 0 | Frameshift deletion | 9.30% (9.20% - 9.40%) |
| chr11.6413139.C.A | p.Pro282Thr | 2.00E-04 | NA | 2 | 0 | 0 | 0 | Damaging | 9.70% (9.70% - 9.70%) |
| chr11.6413142.G.A | p.Ala283Thr | 1.00E-04 | NA | 1 | 0 | 0 | 0 | Damaging | 11.10% (11.00% - 11.20%) |
| chr11.6413181.C.T | p.Arg296Trp | 5.44E-05 | NA | 3 | 0 | 0 | 0 | Damaging | 87.10% (85.50% - 88.60%) |
| chr11.6413187.C.G | p.Leu298Val | NA | NA | 1 | 0 | 0 | 0 | Damaging | 100.90% (99.90% - 101.80%) |
| chr11.6413235.G.A | p.Val314Met | 0 | 0 | 1 | 0 | 0 | 0 | Damaging | 55.30% (54.80% - 55.80%) |
| chr11.6413278.G.C | p.Ser328Thr | NA | NA | 1 | 0 | 0 | 0 | Probably damaging | 34.20% (34.00% - 34.30%) |
| chr11.6413290.C.G | p.Pro332Arg | 5.60E-03 | 1.30E-03 | 69 | 1 | 48 | 0 | Damaging | 30.40% (30.30% - 30.50%) |
| chr11.6413296.T.C | p.Ile334Thr | 0 | 0 | 0 | 0 | 1 | 0 | Damaging | 78.80% (78.40% - 79.10%) |
| chr11.6414486.C.T | p.Arg378Cys | 0 | NA | 1 | 0 | 0 | 0 | Damaging | 37.10% (36.70% - 37.60%) |
| chr11.6414487.G.A | p.Arg378His | 2.00E-04 | 0 | 2 | 0 | 3 | 0 | Damaging | 63.80% (63.60% - 64.00%) |
| chr11.6414492.A.T | p.Ile380Phe | NA | NA | 1 | 0 | 0 | 0 | Damaging | 41.00% (40.40% - 41.50%) |
| chr11.6414543.A.G | p.Asn397Asp | NA | NA | 0 | 0 | 1 | 0 | Probably damaging | 67.80% (67.30% - 68.30%) |
| chr11.6414558.G.A | p.Ala402Thr | 0 | NA | 1 | 0 | 0 | 0 | Damaging | 64.20% (64.00% - 64.40%) |
| chr11.6414877.C.T | p.His432Tyr | 5.44E-05 | NA | 1 | 0 | 0 | 0 | Probably damaging | 111.60% (111.40% - 111.90%) |
| chr11.6414911.G.A | p.Arg443Gln | 4.00E-04 | 6.00E-04 | 5 | 0 | 4 | 0 | Damaging | 24.00% (24.00% - 24.00%) |
| chr11.6415556.T.C | p.Tyr539His | NA | NA | 0 | 0 | 1 | 0 | Damaging | 10.20% (10.20% - 10.20%) |
| chr11.6415622.C.T | p.Arg561Cys | 0 | NA | 0 | 0 | 1 | 0 | Damaging | 47.50% (47.30% - 47.80%) |
| chr11.6415712.C.T | p.Arg591Cys | 0 | 0 | 3 | 0 | 1 | 0 | Damaging | 40.60% (40.50% - 40.60%) |
| chr11.6415730.G.A | p.Ala597Thr | NA | NA | 2 | 0 | 0 | 0 | Probably damaging | 50.80% (50.70% - 51.00%) |
| chr11.6415736.C.T | p.Leu599Phe | NA | NA | 0 | 0 | 1 | 0 | Damaging | 27.90% (27.80% - 28.10%) |
| chr11.6415746.G.A | p.Arg602His | 0 | NA | 1 | 0 | 2 | 0 | Damaging | 29.50% (29.20% - 29.80%) |
| chr11.6415769.C.T | p.Arg610Cys | 0 | 0 | 0 | 0 | 1 | 0 | Damaging | 73.70% (73.50% - 73.80%) |
| Rare benign variants |  |  |  |  |  |  |  |  |  |
| chr11.6411878.G.A | p.Arg17Gln | 1.00E-04 | NA | 3 | 0 | 2 | 0 | NA | 119.60% (118.70% - 120.60%) |
| chr11.6411917.T.G | p.Leu30Arg | NA | NA | 1 | 0 | 1 | 0 | NA | 77.10% (76.80% - 77.50%) |
| chr11.6411958.G.C | p.Ala44Pro | 2.00E-04 | 6.00E-04 | 0 | 0 | 1 | 0 | Benign | 126.80% (126.30% - 127.30%) |
| chr11.6412022.C.T | p.Ser65Phe | NA | NA | 0 | 0 | 1 | 0 | Benign | 99.60% (98.00% - 101.30%) |
| chr11.6412054.A.C | p.Ile76Leu | NA | NA | 1 | 0 | 0 | 0 | Benign | 102.20% (101.40% - 102.90%) |
| chr11.6413089.G.C | p.Gly265Ala | NA | NA | 2 | 0 | 2 | 0 | Benign | 57.10% (57.00% - 57.30%) |
| chr11.6413277.A.G | p.Ser328Gly | NA | NA | 1 | 0 | 0 | 0 | Benign | 82.30% (81.70% - 82.90%) |
| chr11.6413305.A.G | p.Asn337Ser | 5.44E-05 | NA | 2 | 0 | 0 | 0 | Benign | 51.80% (51.50% - 52.00%) |
| chr11.6413377.G.A | p.Arg361His | 5.44E-05 | 0 | 2 | 0 | 1 | 0 | Benign | 68.30% (68.00% - 68.50%) |
| chr11.6415259.G.A | p.Gly492Ser | 2.00E-04 | 6.00E-04 | 0 | 0 | 3 | 0 | Benign | 49.10% (48.60% - 49.60%) |
| chr11.6415583.C.T | p.Pro548Ser | 5.44E-05 | NA | 0 | 0 | 1 | 0 | Benign | 64.80% (64.50% - 65.20%) |
| chr11.6415590.C.T | p.Thr550Ile | NA | NA | 1 | 0 | 0 | 0 | Benign | 60.20% (59.60% - 60.90%) |
| chr11.6415634.G.A | p.Asp565Asn | 1.00E-04 | NA | 1 | 0 | 1 | 0 | Benign | 105.30% (104.90% - 105.70%) |
| chr11.6415639.G.T | p.Met566Ile | 2.00E-04 | NA | 1 | 0 | 3 | 0 | Benign | 70.40% (70.30% - 70.60%) |
| chr11.6415704.C.T | p.Thr588Met | 0.001 | 0.0013 | 9 | 0 | 10 | 0 | Benign | 62.30% (59.30% - 65.50%) |
| Common variants |  |  |  |  |  |  |  |  |  |
| chr11.6415463.G.A | p.Gly508Arg | 0.137 | 0.153 | 1018 | 83 | 1329 | 115 | Damaging | 55.60% (55.40% - 55.80%) |
| chr11.6415539.C.T | p.Pro533Leu | 0.007 | 0.014 | 76 | 1 | 76 | 0 | Damaging | 72.10% (71.90% - 72.30%) |

To confirm the veracity of the *GBA1* and *SMPD1* rare variants identified from whole-exome sequencing, we performed capillary sequencing on 35 rare *SMPD1* variants and 43 *GBA1* variants where DNA was available, with a concordance rate of 100 percent (**Table S4**).

### Validation of the association between rare qualifying variants in SMPD1 and increased susceptibility to Parkinson’s disease

As the association between rare *GBA1* variants and increased risk of Parkinson’s disease has been extensively confirmed in many studies^26^, we focused on evaluating whether the association between rare *SMPD1* variants and Parkinson’s disease could be replicated in additional, independently ascertained samples (**Table S5B**). We pursued this question in a validation analysis comprising 5,585 participants with Parkinson’s disease and 5,642 controls from three studies of Asian and European ancestry (**Table S2B**). Participants from these three studies have previously undergone whole-exome or whole-genome sequencing, with documented equal re-sequencing of cases and controls, and had data on *SMPD1* available. Stratified meta-analysis of the three replication collections showed that carriers of qualifying variants in *SMPD1* remain over-represented in participants with Parkinson’s disease (139 carriers out of 5,585 affected individuals, 2.5%) compared to unaffected control individuals (87 carriers out of 5,642 controls, 1.5%) (OR = 1.45, *P* = 0.0071). When all discovery and replication samples were meta-analyzed (9,883 persons with Parkinson’s disease and 11,154 unaffected individuals across seven independent strata), genome-wide significant association between carriage of *SMPD1* qualifying variants and susceptibility to Parkinson’s disease was observed (OR = 1.77, *P* = 4.94 × 10^-8^, **Figure 1**).

We were struck by the significant heterogeneity (I^2^ _index for heterogeneity_ = 79.6%, *P* = 0.027) in odds ratios between the discovery (OR = 2.32) and replication (OR = 1.45) datasets, and asked whether it could be due to misclassification of qualifying genetic variants by the PolyPhen-2 algorithm. Resolving this would necessitate the design of functional biological assays to measure the activity of each *SMPD1* allele for comparison against wild-type *SMPD1*. Alleles that demonstrate significant loss of function compared to wild-type would thus be included in the revised gene-based burden test. Conversely, alleles that show functional activity that were indistinguishable to wild-type *SMPD1* would be excluded from gene-based burden tests regardless of PolyPhen-2 prediction. *SMPD1* encodes for acid sphingomyelinase (ASM), an enzyme responsible for breaking down sphingomyelin to ceramide and phosphorylcholine ^42^. Complete deficiency of SMPD1 leads to Niemann-Pick’s disease type A/B. We thus designed enzymatic assays to measure lysosomal acid sphingomyelinase enzymatic activity for all 125 *SMPD1* genetic variants that were detected from all (discovery and replication) samples analyzed (**Figure 2A**, **Figure S7**, **Figure S8, Figure S9**, **Table S5B**, **Supplementary Results**).

**Figure 2:**
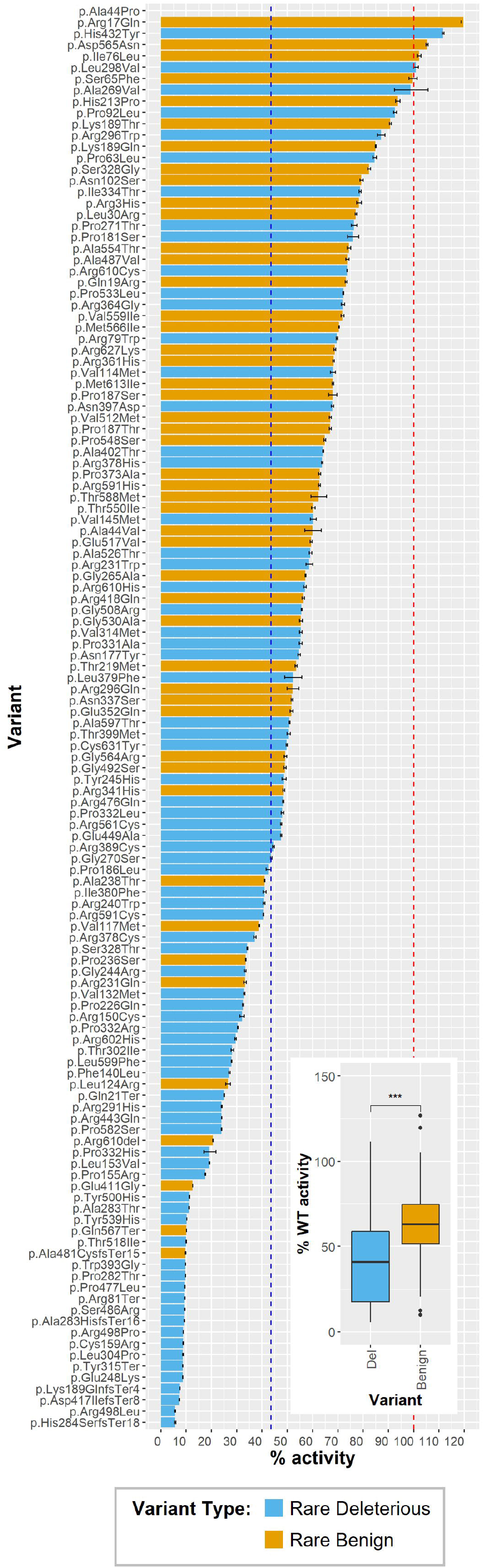
ASM activity levels of *SMPD1* variants in Discovery and Replication datasets. Activity levels presented as percentage of wild-type ASM activity, error bars represent standard deviation. Inset: predicted deleterious variants (blue bars) have significantly lower ASM activity than predicted benign variants (orange bars). Median, upper and lower quartiles showed in boxplot. ***: t-test *P* < 0.001. Red dotted line: 100% activity of wild-type variants. Blue dotted line: activity level (43.58%) significantly associated with variant carrier frequency in case and control. Details in **Table S5B**.

The functional spectrum of acid sphingomyelinase enzymatic activity for all detected *SMPD1* variants ranged from normal (> 90% of wild-type activity) to severe loss-of-function (< 20% of wild-type activity). Two of the variants tested have been previously reported to be causative of Niemann-Pick’s disease type A/B (*SMPD1* p.Leu304Pro [also known as p.Leu302Pro^50^] and p.Arg498Leu^51^) and thus served as positive controls for our experiment. Unsurprisingly, both variants ranked in the bottom 10^th^ percentile of acid sphingomyelinase activity (with p.Leu304Pro showing only 8.7% activity and p.Arg498Leu showing 5.6% activity compared to wild-type SMPD1) in our assay. Across all 125 variants tested, we observed that variants predicted to be damaging by PolyPhen-2/LOFTEE correlated significantly -albeit imperfectly- with lower ASM activity levels compared to variants predicted to be benign (t-test *P* = 1.11 × 10^-5^). We observed that carriage of *SMPD1* variants with less than 44 percent enzymatic activity (translated to 56 percent loss of function or more) show the strongest association with Parkinson’s disease risk in both the discovery cohort (stratified CMH; OR = 2.37, *P* = 4.35 × 10^-7^), replication cohort (stratified CMH; OR = 2.18, *P* = 4.80 × 10^-10^), and the combined discovery and replication cohorts (stratified CMH; OR = 2.24, *P* = 1.25 × 10^-15^, **Figure 3**, **Figure S10**, **Figure S11**, **Figure S12**, **Supplementary Results**).

**Figure 3:**
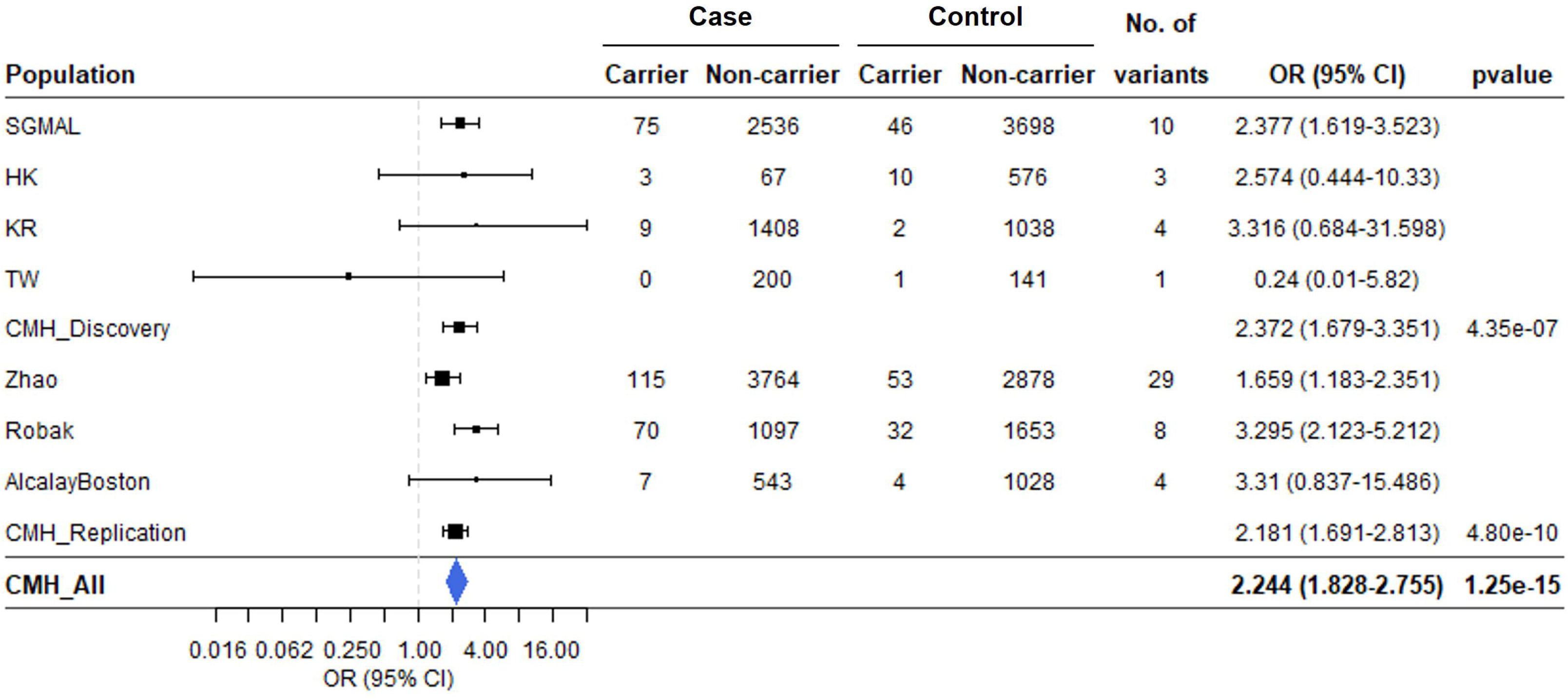
Replication of *SMPD1* association in Chinese and European datasets for rare variants with *≤* 43.58% ASM activity level. Error bars represent 95% confidence interval (CI).

## Discussion

Our observations resulting from whole-exome sequencing, replication in additional independent samples, and functional enzymatic assays suggest that heterozygous carriage of deficient *SMPD1* alleles is associated with 2.2-fold higher odds of Parkinson’s disease. The top two genes emerging from this whole exome sequencing effort, *GBA1* and *SMPD1*, are both involved in lysosomal storage diseases that present early in life (often in childhood) with severe clinical symptoms. Homozygous loss-of-function mutations in *GBA1* causes Gaucher’s disease whereas homozygous loss of SMPD1 function invariably results in Niemann-Pick disease type A/B.

*GBA1* has been extensively evaluated since its identification as a Parkinson’s disease risk gene^26^, with patients carrying rare, protein-altering variants robustly associating with increased risk of Parkinson’s disease, and a more severe clinical phenotype^52^. Conversely, the association between protein-altering *SMPD1* variants and Parkinson’s disease have been inconsistent. The first study linking *SMPD1* with Parkinson’s disease reported that carriage of the p.Leu304Pro variant was associated with 9-fold increased odds of disease^53^. *SMPD1* p.Leu304Pro was a loss-of-function, founder mutation in persons of Ashkenazi Jewish ancestry and was not present for further evaluation in most other ancestry groups^54^. Since then, other smaller studies have shown equivocal evidence in support for other *SMPD1* variants^29,30,53,55,56^ and others have reported no significant association between *SMPD1* variants and susceptibility to Parkinson’s disease^22,57-59^. Similarly, *SMPD1* was not identified in the largest whole exome sequencing study in samples of European ancestry, which otherwise reported strong association at *GBA1* and *LRRK2* (driven by the rare p.Gly2019Ser variant that is not present in Asian populations)^27^. We suspect these inconsistent *SMPD1* reports could be due to the limitations of the bioinformatic algorithms used to predict and identify ‘damaging’ genetic variants. Indeed, incorrect functional classification of genetic variants drastically reduces statistical power for rare-variant burden tests^28^.

By using biological assays based upon the catalytic activity of SMPD1 to classify the functional impact of all 125 *SMPD1* alleles detected in this study, we were able to only include genuine functionally impaired variants in the gene-based burden test. Our experimental assay was adequately controlled as it was able to correctly classify mutations known to cause Niemann-Pick disease type A/B. This is a significant step forward from the current use of predictive bioinformatic algorithms to distinguish variants that impair protein function from ‘benign’ variants. Our data clearly shows that carrying one functionally defective copy of *SMPD1* is sufficient to associate with more than 2-fold increased risk of sporadic, adult-onset Parkinson’s disease. In contrast, complete loss of *SMPD1* causes childhood onset Niemann-Pick disease type A/B. Bearing in mind that the acid sphingomyelinase enzyme encoded by *SMPD1* catalyzes the conversion of sphingomyelin to phosphocholine and ceramide^42^, previous investigators observed significant differences in sphingomyelin and ceramide lipid species between post-mortem brains from patients with Parkinson’s disease compared to brains from unaffected donors^60,61^. Taken together, the spectrum of functional *SMPD1* alleles detected in our large-scale sequencing study affirm a role for acid sphingomyelinase in the pathobiology of neurodegenerative diseases.

### Limitations

Our study has several limitations. First, rare, protein-altering genetic variants in *GBA1* and *SMPD1* still account for a small proportion of Parkinson’s disease patients. Despite the large number of participants that were sequenced, we only observed 8 individuals who carried a protein-altering variant in both *GBA1* and *SMPD1*. Of these, two were Parkinson’s disease patients who carried a known damaging variant in *GBA1* (p.Leu483Pro) and *SMPD1* (p.Pro332Arg). Their variable age of disease onset (at 48 and 62 years of age) is in keeping with the high genetic heterogeneity observed in sporadic Parkinson’s disease. Future work involving more participants carrying mutations in both genes will be needed to resolve the question of possible interaction between both genes. Second, the acid sphingomyelin enzymatic assay only measures the catalytic activity of SMPD1. Thus, genetic variants that result in decreased protein stability would not be accurately evaluated by this assay, as well as variants that result in post-translational mis-localization of SMPD1. Third, the concordance between our *in-vitro* acid sphingomyelinase enzymatic activity assays and previously reported measurements based on dried blood spots was approximately 50 percent^31^. This discordance could reflect variability in laboratory techniques as well as opportunities to perform experiments in multiple biological replicates for more accurate ascertainment of enzymatic activity. Reassuringly, one such discordant variant (*SMPD1* p.Ala487Val) was re-assayed by a third party, and their conclusions agreed with our findings that this variant does not qualify as functionally deficient^62^. Finally, genes with variants of similar effect sizes to *SMPD1*, but present at lower combined carrier frequencies in Asian populations may be missed in our discovery study (**Figure S13**). Larger multi-center studies will facilitate the discovery of additional genes with protein-altering variants influencing PD risk.

### Conclusion

Our study suggests that rare independent alleles in *SMPD1* that impair acid sphingomyelinase activity are associated with increased risk of Parkinson’s disease. These observations were made across multiple case-control collections (N = 21,037 participants, of which 9,883 had Parkinson’s disease) that were ascertained and sequenced independently from one another at different times and under different experimental conditions. Our findings will inform further work on understanding the specific aspects of impaired sphingolipid and ceramide metabolism in the pathogenesis of Parkinson’s disease.

## Supporting information

Supplementary Methods, Results and Figures

## Data availability

Summary statistics, acid sphingomyelinase enzymatic activity assay raw results and Western blots are available upon request. Individual level genotype data are not publicly available because of consent and privacy issues but will be made available upon reasonable request and following approval by the relevant institutional review board.

## Code availability

The codes used in this study can be found on GitHub (https://github.com/fjnlab/PD_exome-seq). Software and software packages used include: R (version 4.0.2) and R packages data.table (version 1.14.0), dplyr (version 1.0.5), tidyr (version 1.1.2), ggplot2 (version 3.3.6), forestplot (version 2.0.1); GATK (version 3.7); Picard (version 2.21.7); VCFtools (version 0.1.16); BCFtools (version 1.9); PLINK (version 1.9); Fratools PERL package (https://github.com/atks/fratools); Ensembl Variant Effect Predictor (VEP) software (version 104); VEP plugin LOFTEE (version 1.0.3); PolyPhen-2 (version 2.2.3r406); dbNSFP (version 3.5a).

## Acknowledgements

We thank Wee-Yang Meah (Genome Institute of Singapore), Shin Hui Ng (National Neuroscience Institute), Wei Ling Beh, Winnie Tay, Ken Wong and Andrew Ang (Nanyang Technological University Singapore) for administrative support. We thank Su Wei Lim and Yun Wutt Yi Oo for assistance in structural modeling. This work is supported by the Singapore Ministry of Health’s National Medical Research Council Open Fund Large Collaborative Grant (MOH-000207; to E.-K.T.) Open Fund Individual Research Grant (MOH-000559; to J.N.F.; MOH-001110; to Z.W.), Singapore Translational Research Investigator Award (STaR) (MOH-000435; to. T.A. and Z.W.), as well as the Singapore Ministry of Education Academic Research Fund Tier 2 (MOE-T2EP30220-0005; to J.N.F.) and Tier 3 (MOE-MOET32020-0004; to J.N.F.). C.C.K is supported by the Singapore National Research Foundation Investigatorship (NRF-NRFI2018-01). S.-Y.L. and A.-H.T. are supported by the University of Malaya Parkinson’s Disease and Movement Disorders Research Program (PV035-2017).

## Notes

### Competing Interest Statement

The authors have declared no competing interest.

### Author Declarations

This study was approved by the ethics committees or institutional review boards of the respective institutions (SingHealth Centralized Institutional Review Board CIRB 2002/008/A and 2019/2334 and Nanyang Technological University Institutional Review Board IRB-2016-08-011).

