## Supplementary Methods, Results and Figures for "Exome sequencing in Asian populations identifies rare deficient *SMPD1* alleles that increase risk of Parkinson’s disease"

### Supplemental Data

### Inventory of supporting information

*List of Supplemental Tables*

**Table S1**: Discovery samples numbers before and after quality control

**Table S2A**: Demographics of Discovery samples included in final analysis

**Table S2B**: Sample details of Replication dataset

**Table S3**: Summary of sample quality control conducted

**Table S4A**: Sanger validation PCR conditions and results for *SMPD1* variants

**Table S4B**: Sanger validation PCR cycling conditions

**Table S5A:** *SMPD1* variant-specific cloning primer sequences.

**Table S5B:** *SMPD1* variants in Discovery and Replication datasets

**Table S6A:** *GBA* rare variants from the Discovery dataset

**Table S6B:** *SMPD1* rare synonymous variants from the Discovery dataset

**Table S7A:** Association testing of familial Parkinson's disease related genes in Discovery dataset

**Table S7B:** Association testing of lysosomal storage disease related genes in Discovery dataset

*List of Supplemental Figures*

**Figure S1:** Variant calling and filtering pipeline for Discovery dataset.

**Figure S2**: Ancestry principal components analysis of samples that passed quality control.

**Figure S3:** Assessment of reproducibility of the ASM enzymatic activity assay.

**Figure S4**: Replication of *GBA* association in Chinese and European datasets based on rare predicted pathogenic variants

**Figure S5**: Onset ages of PD cases that carry rare deleterious variants in (A) *GBA*, (B) *SMPD1*, and (C) *GBA* and *SMPD1*.

**Figure S6:** QQplot of all autosomal genes with OR > 1 from the discovery dataset.

**Figure S7**: Wildtype SMPD1 protein expression and variant-associated truncated protein expression in HEK293 cells.

**Figure S8**: In silico structural analysis of p.Pro282Thr, p.Tyr500His and p.Asn177Tyr mutations in SMPD1.

**Figure S9:** Western blots of SMPD1 protein expression for all 125 assayed variants.

**Figure S10:** Iterative linear discriminant analysis of *SMPD1* variants defined as functionally deficient at various ASM enzymatic activity thresholds.

**Figure S11**: Distribution of ASM enzymatic activity levels of rare *SMPD1* variants in relation to its (A) effects on tertiary structure integrity and (B) localization to protein domains.

**Figure S12:** Forest plot of rare *SMPD1* variants from Discovery and Replication datasets that (A) affect ASM tertiary structure, (B) localized to the signaling peptide, (C) localized to the signaling peptide, (D) localized to the proline-rich linker, (E) localized to the catalytic metallophosphate domain, and (F) localized to the C terminus.

**Figure S13**: Statistical power to identify PD-associated genes at exome-wide significance.

### Supplemental Methods

*Cloning of SMPD1 alleles for acid sphingomyelinase enzymatic activity assays*

Fragments containing *SMPD1* alleles were generated by one of the three approaches depending on the proximity of the *SMPD1* variant to the 3' or 5' ends of the *SMPD1* open reading frame. The first approach involved generating *SMPD1* allele from two separate overlapping fragments which contained the variant-specific mutation. These two fragments were amplified from the open reading frame of the *SMPD1* canonical transcript ENST00000342245.4 using primer pairs detailed in **Table S5A**. One fragment will be generated from amplification with the forward end primer (SMPD1_FWD) and variant-specific reverse primer, while the other fragment will be generated from amplification with the variant-specific forward primer and reverse end primer (SMPD1_REV). Both fragments and the linearized pLVX-CMV vector will then be joined by Gibson assembly (# E2621S, New England Biolabs). The second approach involved generating a single fragment for *SMPD1* alleles close to the 3' or 5' ends of the *SMPD1* open reading frame (p.Arg3His, p.Arg17Gln, p.Arg627Lys, p.Cys631Tyr) by incorporating the variant-specific mutation into the variant-specific primer to produce a single full length fragment following PCR with SMPD1_REV or SMPD1_FWD. The single fragment and the linearized pLVX-CMV vector will then be joined by Gibson assembly (#E2621S, New England Biolabs). In the third approach, fragments for *SMPD1* alleles (p.Ala283HisfsTer16, p.His284SerfsTer18, p.Leu304Pro, p.His432Tyr, p.Gly492Ser, p.Arg498Leu) that were technically challenging to generate with the PCR-based approach described above were synthesized using the GenBrick synthesis service from GenScript, before restriction digestion and ligation with the linearized pLVX-CMV vector. Sanger sequencing was conducted on all generated plasmids to confirm variant sequences before transduction into HEK 293 cells for the acid sphingomyelinase enzymatic activity assay.

*In silico structural analysis of p.Pro282Thr, p.Tyr500His and p.Asn177Tyr variants*

Steric clashes due to p.Pro282Thr, p.Tyr500His and p.Asn177Tyr mutations were evaluated using the mutagenesis toolbox in PyMol version 2.5.2 based on the atomic coordinates of human aSMase bounded with zinc and phosphocholine (Protein Database Bank ID: 5I85, version 2.1) [1].

### Supplemental Results

*Lead variants contributing to SMPD1 association to Parkinson's disease in the Discovery and Replication datasets.*

We observed that the association between rare qualifying variants in *SMPD1* and increased risk of Parkinson's disease was driven by one particular variant (p.Pro332Arg) in the Discovery dataset (allele frequency in participants with Parkinson's disease = 0.8 percent, allele frequency in unaffected controls = 0.4 percent, odds ratio (OR) = 2.39, $P = 5.01 \times 10^{-6}$). This particular variant was also the lead variant in Chinese replication dataset [2] (allele frequency in participants with Parkinson's disease = 1%, allele frequency in unaffected control individuals = 0.53%; OR = 1.95, $P = 0.00132$). When we excluded p.Pro332Arg from the analysis, a significant association between burden of rare qualifying variants in *SMPD1* and increased risk of Parkinson's disease remained (OR = 2.31, $P = 0.00369$), suggesting that the association between rare, independent genetic variants in *SMPD1* was not confined to p.Pro332Arg alone.

*SMPD1* encodes for acid sphingomyelinase (ASM). Analysis of the crystal structure of ASM predicted that the p.Pro332Arg alteration would result in a breakage of hydrophobic contacts with the p.Ala402 side chain. This in turn exposes a hydrophobic pocket and might affect the folding of the ASM tertiary structure [1]. Such functional predictions based on protein crystallography is fully congruent with our observation that the *SMPD1* p.Pro332Arg alteration results in an ASM protein with enzymatic activity of only 30.4% compared to wild-type ASM (**Table 1, Table S5B**).

Both our data and public databases such as gnomAD suggest that *SMPD1* p.Pro332Arg is likely to be an Asian specific variant that was also observed in the Chinese Replication dataset [2] but not in the European Replication datasets [3,4]. At the same variant position, the c.995C>A mutation results in p.Pro332His amino acid substitution that was reported in one Parkinson's disease case from Chinese replication dataset [2], while the c.995C>T mutation results in p.Pro332Leu that was reported in one Parkinson's disease case from the European Replication dataset [4]. In contrast, no unaffected control individuals carried either the p.Pro332His or p.Pro332Leu substitution. All three *SMPD1* variants (p.Pro332Leu, p.Pro332His, and p.Pro332Arg) have ASM enzymatic activity <50% compared to wild-type SMPD1 and their enrichment in persons with Parkinson's disease compared to unaffected controls suggest them to be pathogenic alterations.

The lead variant for European samples, *SMPD1* p.Glu411Gly, was only reported in the Robak dataset [4] where it was significantly enriched in Parkinson's disease cases as compared to controls ($P$ = 8.99 x $10^{-11}$, OR = 4.83) although predicted by PolyPhen-2 to be benign. However, biological testing revealed this variant to have severe loss-of-function (with only 12.5 percent of enzymatic activity remaining) such that its inclusion into the functionally classified gene-based burden test resulted in a marked increase in the odds ratios. Further assessment of this p.Glu411Gly variant in additional independent samples would be needed to affirm its role in Parkinson's disease, as it was not detected in the other US-based replication cohort [3].

*Validation of the association between rare qualifying variants in GBA and increased susceptibility to Parkinson's disease.*

We attempted to evaluate whether carriage of qualifying variants in *GBA* was associated with increased risk of Parkinson's disease in persons of Chinese ancestry. This was pursued using published *GBA* data [2] after variant re-annotation and re-filtering using the same criteria that was applied to the discovery dataset. We observed that persons carrying rare, protein-altering *GBA* variants were associated with a 9-fold increased odds of Parkinson's disease in this Chinese replication dataset alone (with 205 out of 3,879 persons [5.28%] with Parkinson's disease carrying *GBA* rare variants compared to 18 out of 2,931 unaffected controls [0.61%] carrying *GBA* rare variants; $P$ = 7.9 × $10^{-27}$). This association remained robustly significant upon meta-analysis of all participants of East Asian ancestry from the discovery and replication collections (OR =

9.53, $P = 8.7 \times 10^{-50}$), as well as when replication samples comprising participants of European ancestry [4] were included ($P = 2 \times 10^{-59}$, OR = 7.22; **Figure S3**).

*SMPD1 protein expression in HEK293-based ASM activity assay model*

Western blotting conducted with anti-SMPD1 antibody confirmed that there was no native SMPD1 expression in the HEK293 cells used in the assay. There was no observed SMPD1 expression when an empty pLVX-CMX vector was transduced into the HEK293 cells (**Figure S7A**).

Out of the 125 variants transduced into the HEK293 cells and tested, eight variants resulted in the translation of smaller SMPD1 proteins as compared to wild type SMPD1 variant (**Figure S7B**). These variants include three predicted damaging missense mutations (p.Asn177Tyr, p.Pro282Thr, p.Tyr500His), one predicted pathogenic stopgain variant (p.Tyr315Ter), three predicted deleterious frameshift variants (p.Ala283HisfsTer16, p.His284SerfsTer18, p.Asp417IlefsTer8) and one predicted benign frameshift variant (p.Ala481CysfsTer15; low confidence prediction by LOFTEE). p.Asn177Tyr is localised to the proline-rich linker, while the other seven variants are localized to the metallophosphate domain. All seven variants localised to the metalloproteinase domain have been found to result in low functional activity between 5.6 - 11.2 % as compared to wildtype SMPD1 variant, and have been classified as loss-of-function variants. While p.Asn177Tyr present at the proline-rich linker has a functional score of 54.7% and is unlikely to affect SMPD1 activity.

Missense variant p.Pro282Thr was present in two Parkinson's disease patients from the discovery samples, was absent in all unaffected control individuals, and was unexpectedly found to result in two polypeptide forms of 22 kDa and 26 kDa size. We repeated p.Pro282Thr mutagenesis, cloning and the associated assay multiple times and obtained the same finding of multiple polypeptide sizes and low functional activity from p.Pro282Thr. We hypothesize that the p.Pro282Thr variant disrupt the ASM protein structure which caused potential protein misfolding and eventual degradation by the endoplasmic reticulum-associated protein degradation that generated the two polypeptide forms of 22 kDa and 26 kDa. Missense mutation p.Tyr500His was present only in Parkinson's disease patients of Chinese ancestry [2] and resulted in two polypeptide forms, one similar in size to wildtype SMPD1 (67 kDa) and another slightly smaller polypeptide of approximately 61 kDa. The smaller 61 kDa polypeptide was the more abundantly expressed species (expressed 1.67-fold more than wildtype SMPD1 polypeptide). Missense variant p.Asn177Tyr was present in one Parkinson's disease patients and two controls from the replication Chinese samples [2], similarly resulted in a smaller polypeptide of 61 kDa. The smaller polypeptides observed in p.Tyr500His and p.Asn177Tyr are likely attributed to affected protein processing, glycosylation or maturation. *In silico* structural analysis shows potential loss of interactions for p.Pro282Thr and p.Tyr500His with side chains of surrounding structures which may affect protein stability (**Figure S8**). Future work will be needed to evaluate whether the low enzymatic activity (relative to wild-type) observed with p.Tyr500His, p.Pro282Thr and p.Asn177Tyr were due to adverse effects on post-translational modification or maturation.

Stopgain variant p.Tyr315Ter that is present only in one Parkinson's disease case from the Chinese replication samples [2] resulted in the formation of a truncated polypeptide that is 33 kDa which is non-functional (functional activity = 8.6%).

Discovery frameshift deletion p.Ala283HisfsTer16, present only in one unaffected individual resulted in the faint expression of a 29 kDa polypeptide. While another Discovery frameshift deletion p.His284SerfsTer18, present in two Parkinson's disease cases, resulted in the expression of two polypeptides with molecular weight of 31 kDa and 34 kDa. Frameshift deletion p.Asp417IlefsTer8 is present in one unaffected individual from the Chinese replication samples [2] and resulted in the expression of a 48 kDa polypeptide. Predicted benign frameshift insertion p.Ala481CysfsTer15 is present in one Parkinson's disease case from the Chinese replication samples [2] and resulted in a 49 kDa polypeptide.

Focus in the future should be placed on variants such as p.Pro282Thr, p.Tyr500His, p.Tyr315Ter, p.His284SerfsTer18 and p.Ala481CysfsTer15 which are only present in cases and absent in unaffected control individuals.

*Alleles with reduced ASM activity are associated with PD in Asian and European populations*

To determine the specific ASM enzymatic activity level threshold that defines functionally deficient *SMPD1* alleles, we use iterative linear discriminant analysis to assess ASM activity levels to obtain maximum discrimination between participants with Parkinson's disease and unaffected control individuals. The level of discrimination is expressed in terms of the odds ratio for burden of enzymatically deficient *SMPD1* variants in both the discovery and replication datasets. This odds ratio is accompanied by a *P*-value for association between burden of enzymatically deficient *SMPD1* variants and risk of Parkinson's disease.

After iterative analysis with enzymatic activity thresholds set from 1% to 100% (relative to wild-type SMPD1) on a nominal scale, we observed that the most significant *P*-value was observed at the threshold of ≤ 43.58% enzymatic activity relative to wildtype (**Figure S10**). At this threshold, carriage of enzymatically deficient *SMPD1* alleles were consistently associated with a significantly increased risk of Parkinson's disease in the discovery collections (OR = 2.37, $P = 4.35 \times 10^{-7}$), replication datasets (OR = 2.18, $P = 4.80 \times 10^{-10}$), and on analysis of all samples evaluated in this report (OR = 2.24, $P = 1.25 \times 10^{-15}$).

We next conducted stratified analysis of the enzymatically deficient *SMPD1* variants by ancestry. We observed that functionally deficient variants present only in Asians have lower average functional score (56 variants, functional score average = 28.6%, range = 5.6% - 59.1%) compared to variants present only in Europeans (18 variants, functional score average = 33.7.6%, range = 5.6% - 55.4%) and variants present in both Asian and European samples (6 variants, functional score average = 52.3%, range = 47.5% - 57.0%).

Lower onset age in carriers of catalytic deficient variants

We seek to determine if carriers of *SMPD1* or *GBA* variants from our Discovery samples had earlier disease onset, defined by earlier age of Parkinson's disease diagnosis, compared to non-carriers. While carriers of *GBA* rare predicted deleterious variants were diagnosed with Parkinson's disease almost 7 years younger than non-carriers (carriers: median = 54.7 years of age, range = 29 - 78 years of age; non-carriers: median = 61.4 years of age, range = 22 - 92 years of age; t-test $P = 3.32 \times 10^{-11}$), the effect was much smaller for carriers of rare predicted deleterious variants at *SMPD1* (carriers: median = 60.2 years of age, range = 32 - 86 years of age; non-carriers: median = 61.2 years of age, range = 22 - 92 years of age; t-test $P = 0.426$), as well as carriers of functionally deficient *SMPD1* (defined as variants with activity levels ≤ 43.58%; carriers: median = 60 years of age, range = 32 - 86 years of age; non-carriers: median = 62 years of age, range = 22 - 92 years of age; t-test $P = 0.216$).

*Distribution of SMPD1 rare variants across the 3-dimensional ASM structure*

We mapped rare *SMPD1* mutations from the Discovery and Replication datasets on to protein domains and 3D human ASM structure (PDB 5i81) and found that majority (65.4%, 51 out of 78) of these mutations were located in the catalytic metallophosphate domain. In contrast, mutations were present in the signaling peptide, saposin, proline-rich linker, and C-terminal domains at a lower rate of 5.1%, 11.5%, 6.4% and 10.3%, respectively. We observed that variants in the metalloproteinase domain (catalytic site) and the saponin domains have the lowest activity out of all domains (**Figure S11**). Furthermore, variants that affect ASM tertiary protein structure integrity [1,5] have significantly lower activity levels (average activity = 30.49%) than variants which do not (average activity = 54.34%, T-test $P = 1.39 \times 10^{-4}$, **Figure S11**). As expected, variants localised to the catalytic metalloproteinase domain were found to be associated with disease status across samples from the discovery and replication datasets (OR = 1.87, 95% CI = 1.57 - 2.24, $P = 1.78 \times 10^{-12}$, **Figure S12**). Variants localizing to the other domains, such as the saponin domain (OR = 0.997, 95% CI = 0.52 -1.91, $P$ = 0.993) did not display similar association with disease status (**Figure S12**).

Two rare deleterious mutations (p.Asn397Asp, p.Asn177Tyr) localized to glycosylated asparagine residues, however their enrichment in controls over cases suggest that glycosylation at these residues are unlikely to be associated with Parkinson's disease. Furthermore, the loss of glycosylation Asn residues at these two positions do not impact ASM activity severely as observed by the moderate ASM functional scores of 67.8% and 54.7% measured for p.Asn397Asp, p.Asn177Tyr.

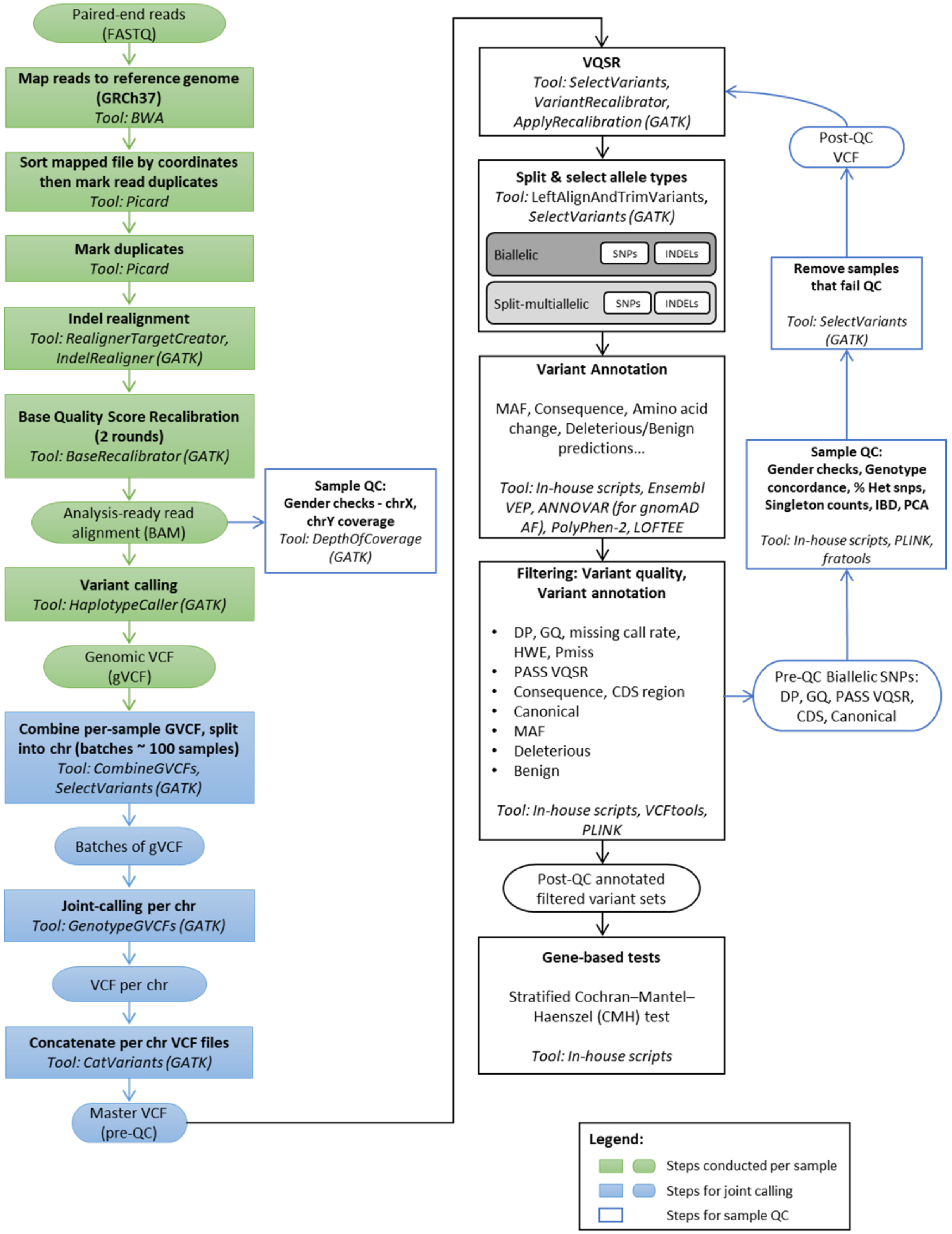


**Figure S1:** Variant calling and filtering pipeline for Discovery dataset.

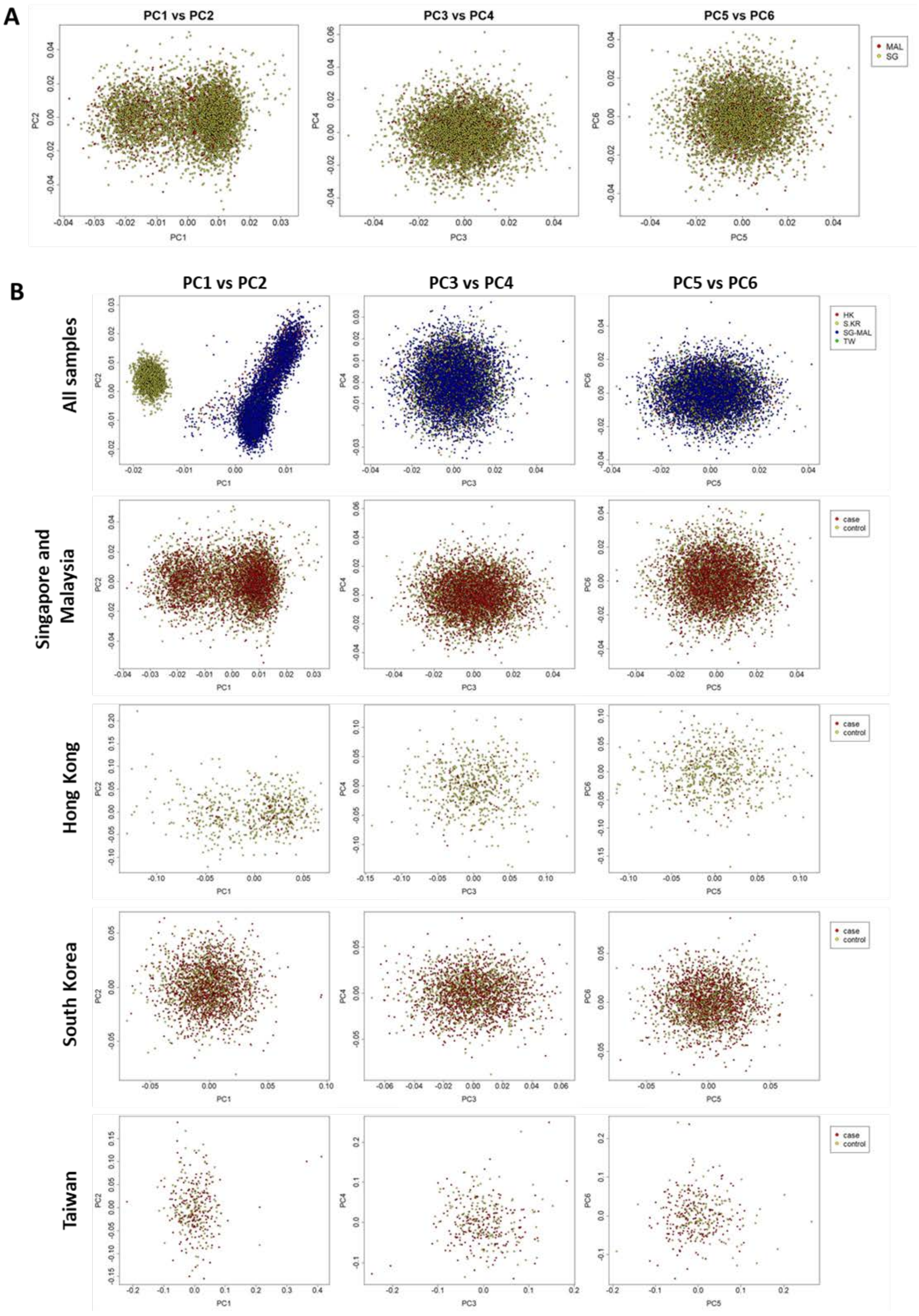


**Figure S2: Ancestry principal components analysis of samples that passed quality control.** (A) Samples from Singapore and Malaysia are similar in genetic background and were considered as one strata. (B) Principal components analysis of all samples and each strata.

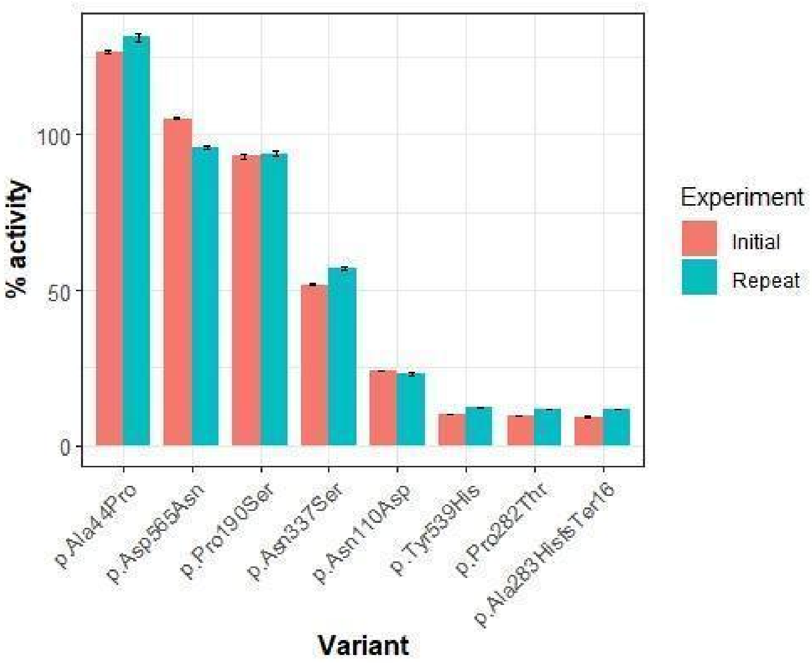


**Figure S3: Assessment of reproducibility of the ASM enzymatic activity assay.** Eight randomly selected *SMPD1* variants were retested by having their cell-lines thawed out, re-passaged, and enzymatic activity re-measured independently in triplicate (shows as vertical bars labeled as 'Repeat'). They were then compared side-by-side with their initial ASM enzymatic activity measurements (shown as vertical bars labeled as 'Initial').

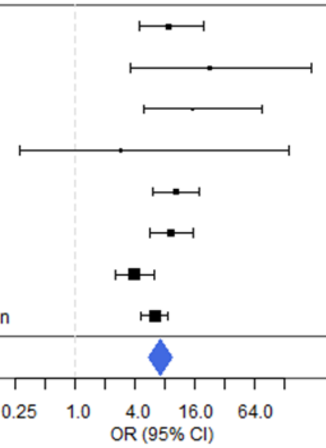


| Population | Case: Carrier | Case: Non-carrier | Control: Carrier | Control: Non-carrier | No. of variants | OR (95% CI) | pvalue |
|---|---|---|---|---|---|---|---|
| SGMAL | 60 | 2551 | 10 | 3734 | 13 | 8.779 (4.449-19.28) | |
| HK | 5 | 65 | 2 | 584 | 2 | 22.227 (3.555-237.031) | |
| KR | 59 | 1358 | 3 | 1037 | 18 | 15.009 (4.87-75.057) | |
| TW | 4 | 196 | 1 | 141 | 3 | 2.87 (0.28-142.606) | |
| CMH_Discovery | | | | | | 10.283 (5.957-17.748) | 4.07e-25 |
| Zhao | 205 | 3674 | 18 | 2913 | 71 | 9.026 (5.554-15.577) | |
| Robak | 78 | 1089 | 30 | 1655 | 27 | 3.949 (2.542-6.28) | |
| CMH_Replication | | | | | | 6.281 (4.567-8.639) | 1.04e-36 |
| **CMH_All** | | | | | | **7.215 (5.484-9.492)** | **1.95e-59** |

**Figure S4: Replication of *GBA* association in Chinese and European datasets based on rare predicted pathogenic variants.**

**A** **GBA**

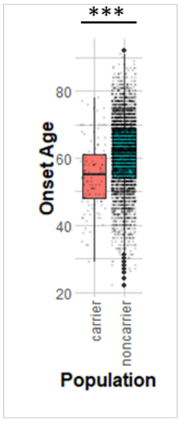


| | | n | mean | sd | median | min | max | ttest pval |
|---|---|---|---|---|---|---|---|---|
| SGMAL | carrier | 55 | 57.7 | 10.9 | 58.0 | 34.0 | 78.0 | 0.001 |
| | non-carrier | 2406 | 62.7 | 10.5 | 64.0 | 22.0 | 92.0 | |
| HK | carrier | 5 | 50.6 | 4.7 | 51.0 | 46.0 | 58.0 | 0.030 |
| | non-carrier | 64 | 57.2 | 10.7 | 58.0 | 24.0 | 78.0 | |
| KR | carrier | 59 | 51.9 | 8.7 | 52.0 | 29.0 | 74.0 | 8.38E-07 |
| | non-carrier | 1353 | 58.3 | 10.1 | 59.0 | 22.0 | 88.0 | |
| TW | carrier | 4 | 58.8 | 9.2 | 60.5 | 47.0 | 67.0 | 0.157 |
| | non-carrier | 196 | 67.3 | 10.5 | 69.5 | 29.0 | 91.0 | |
| All | carrier | 123 | 54.7 | 10.0 | 55.0 | 29.0 | 78.0 | 3.32E-11 |
| | non-carrier | 4019 | 61.4 | 10.7 | 62.0 | 22.0 | 92.0 | |

**B** **SMPD1**

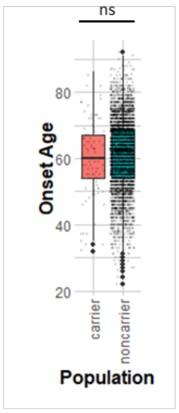


| | | n | mean | sd | median | min | max | ttest pval |
|---|---|---|---|---|---|---|---|---|
| SGMAL | carrier | 78 | 61.3 | 11.4 | 60.5 | 32.0 | 86.0 | 0.295 |
| | non-carrier | 2383 | 62.6 | 10.5 | 63.4 | 22.0 | 92.0 | |
| HK | carrier | 3 | 42.7 | 6.7 | 46.0 | 35.0 | 47.0 | 0.050 |
| | non-carrier | 66 | 57.4 | 10.3 | 58.0 | 24.0 | 78.0 | |
| KR | carrier | 12 | 57.8 | 12.0 | 60.0 | 37.0 | 73.0 | 0.932 |
| | non-carrier | 1400 | 58.1 | 10.2 | 59.0 | 22.0 | 88.0 | |
| TW | carrier | NA | NA | NA | NA | NA | NA | NA |
| | non-carrier | 200 | 67.2 | 10.5 | 69.0 | 29.0 | 91.0 | |
| All | carrier | 93 | 60.2 | 11.8 | 60.0 | 32.0 | 86.0 | 0.426 |
| | non-carrier | 4049 | 61.2 | 10.7 | 62.0 | 22.0 | 92.0 | |

**C** **GBA + SMPD1**

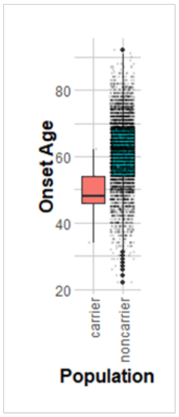


| | | n | mean | sd | median | min | max | ttest pval |
|---|---|---|---|---|---|---|---|---|
| SGMAL | carrier | 3 | 48.0 | 14.0 | 48.0 | 34.0 | 62.0 | 0.212 |
| | non-carrier | 2458 | 62.6 | 10.6 | 63.0 | 22.0 | 92.0 | |
| HK | carrier | 1 | 46.0 | NA | 46.0 | 46.0 | 46.0 | NA |
| | non-carrier | 68 | 56.9 | 10.5 | 57.5 | 24.0 | 78.0 | |
| KR | carrier | 1 | 54.0 | NA | 54.0 | 54.0 | 54.0 | NA |
| | non-carrier | 1411 | 58.1 | 10.2 | 59.0 | 22.0 | 88.0 | |
| TW | carrier | NA | NA | NA | NA | NA | NA | NA |
| | non-carrier | 200 | 67.2 | 10.5 | 69.0 | 29.0 | 91.0 | |
| All | carrier | 5 | 48.8 | 10.4 | 48.0 | 34.0 | 62.0 | 0.055 |
| | non-carrier | 4137 | 61.2 | 10.7 | 62.0 | 22.0 | 92.0 | |

**Figure S5: Onset ages of Parkinson's disease cases that carry rare deleterious variants in (A) *GBA*, (B) *SMPD1*, and (C) both *GBA* and *SMPD1*.** *** indicates t-test *P* < 0.001.

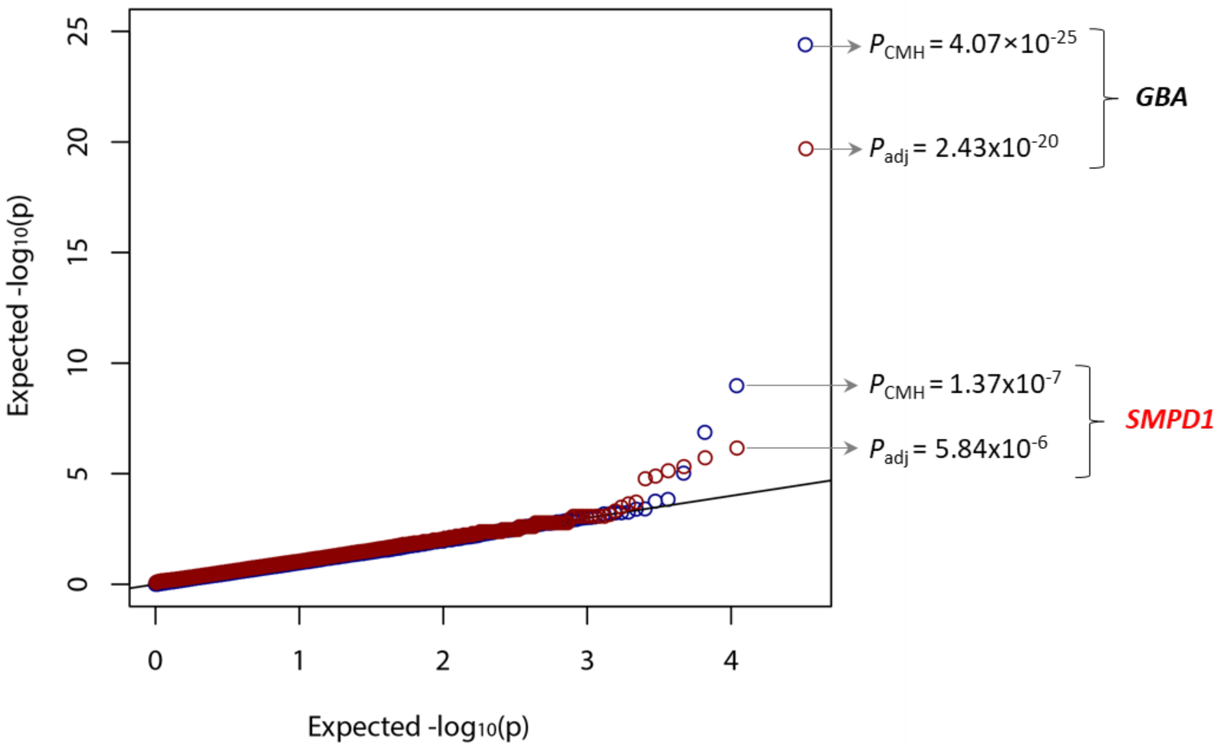


**Figure S6: Quantile-Quantile plot of all autosomal genes with OR > 1 from the discovery dataset.** A stratified CMH test was used to evaluate gene-based burden across the exomes of the study participants from the 5 countries studied ("$P_{CMH}$") and indicated by blue circles. Gene-based burden after adjustment for principal components (PC1-PC3), per sample variant counts and average per sample coverage ("$P_{adj}$") indicated by red circles. Details in **Table S6**.

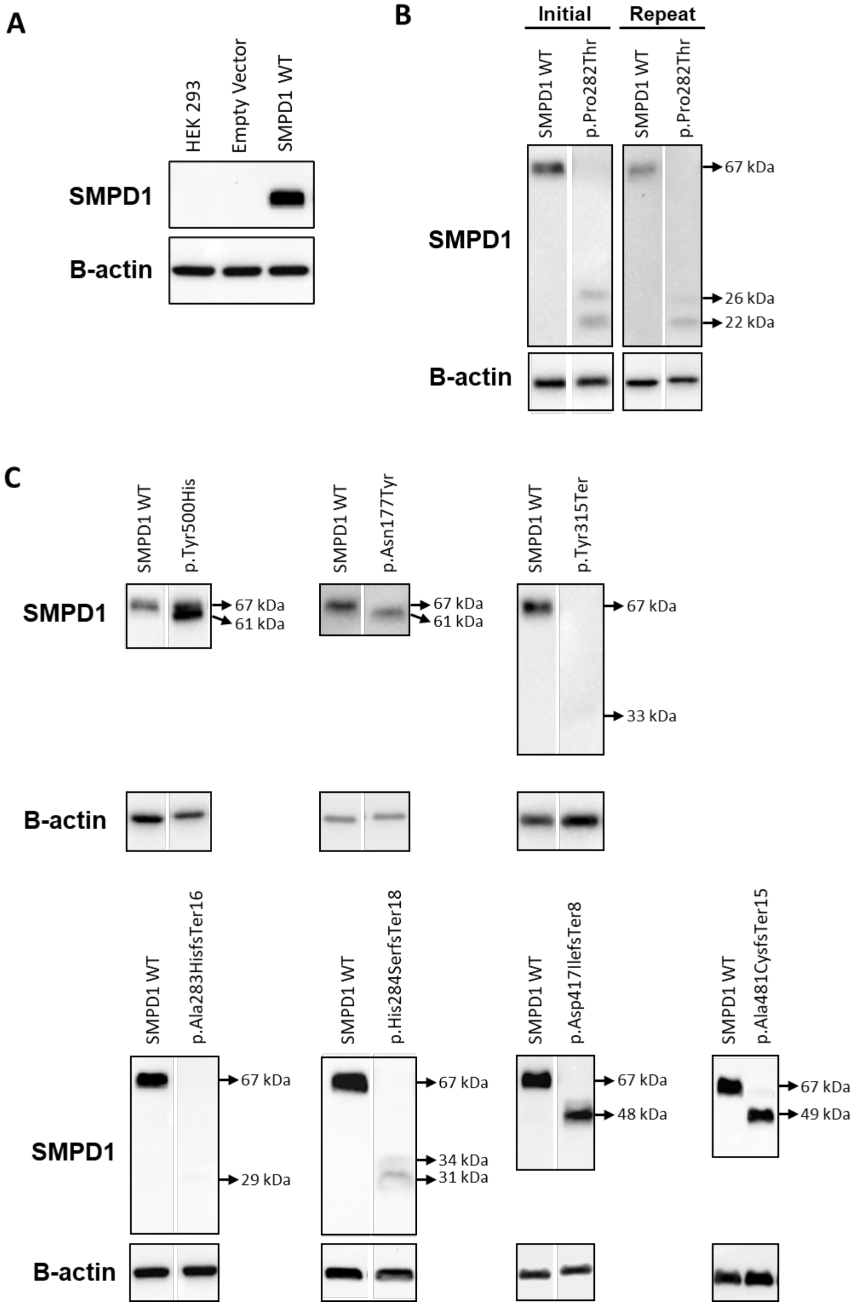


**Figure S7: Wildtype SMPD1 protein expression and variant-associated truncated protein expression in HEK293 cells.** (A) No native SMPD1 expression in HEK293 cells used in the assay (Lane 1). Wild type SMPD1 protein of 67 kDa is observed from HEK293 cells expressing wildtype *SMPD1* cDNA sequence (Lane 3). (B) Two polypeptide sizes (22 kDa and 26 kDa) that were smaller than wildtype SMPD1 polypeptide length of 67 kDa observed from p.Pro282Thr in repeated experiments. (C) Seven other variants result in polypeptide sizes that were smaller than the wildtype SMPD1 polypeptide length of 67 kDa.

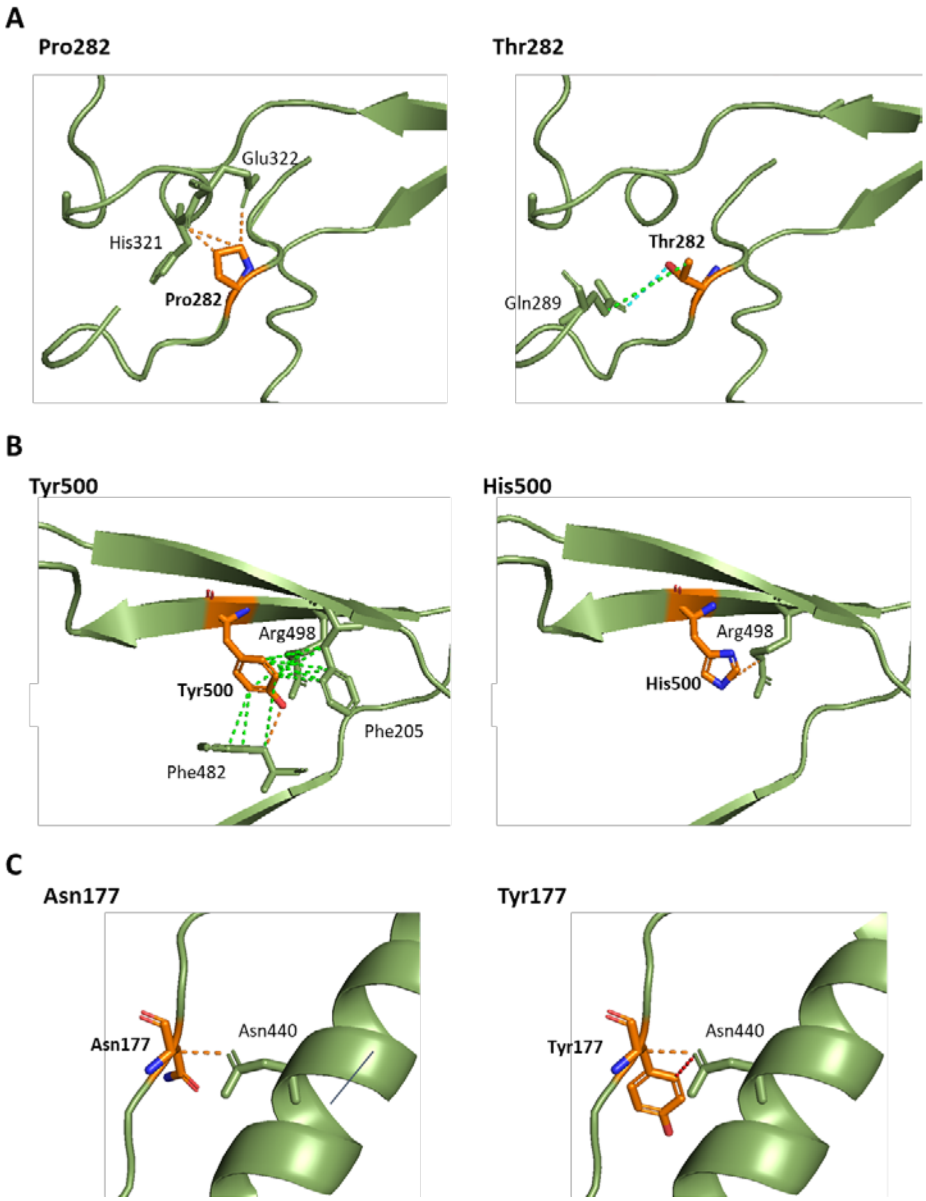


**Figure S8: *In silico* structural analysis of p.Pro282Thr, p.Tyr500His and p.Asn177Tyr mutations in SMPD1.** (A) p.Pro282Thr may lead to loss of polar interactions between Thr282 with side chains at beta3-alpha3 linker region (His321 and Glu322) and a gain in Van der Waals' and hydrophobic interactions with Gln289 at the alpha2-beta2 linker region. (B) p.Tyr500His may result in complete loss of hydrophobic interactions with Phe482 at beta9. (C) p.Asn177Tyr only result in the formation of an additional hydrogen bond between Tyr177 and Asn440. *In silico* analysis conducted based on protein crystallography of ASM tertiary structure 5i85 [1]. Hydrogen bonds (red), hydrophobic interactions (green) and polar bonds (orange).

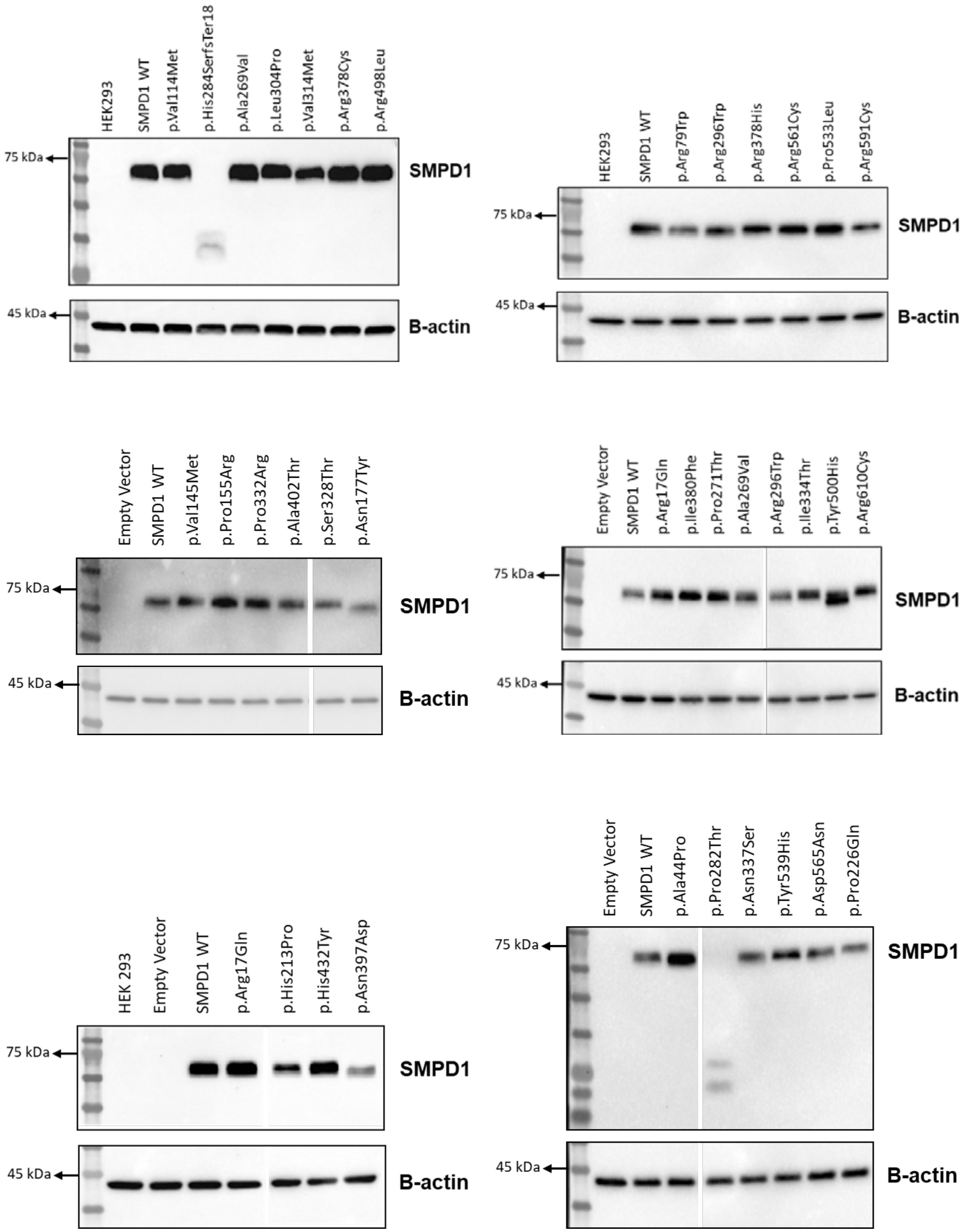

HEK293
SMPD1 WT
p.Val114Met
p.His284SerfsTer18
p.Ala269Val
p.Leu304Pro
p.Val314Met
p.Arg378Cys
p.Arg498Leu
75 kDa
SMPD1
45 kDa
B-actin
HEK293
SMPD1 WT
p.Arg79Trp
p.Arg296Trp
p.Arg378His
p.Arg561Cys
p.Pro533Leu
p.Arg591Cys
75 kDa
SMPD1
45 kDa
B-actin
Empty Vector
SMPD1 WT
p.Val145Met
p.Pro155Arg
p.Pro332Arg
p.Ala402Thr
p.Ser328Thr
p.Asn177Tyr
75 kDa
SMPD1
45 kDa
B-actin
Empty Vector
SMPD1 WT
p.Arg17Gln
p.Ile380Phe
p.Pro271Thr
p.Ala269Val
p.Arg296Trp
p.Ile334Thr
p.Tyr500His
p.Arg610Cys
75 kDa
SMPD1
45 kDa
B-actin
HEK 293
Empty Vector
SMPD1 WT
p.Arg17Gln
p.His213Pro
p.His432Tyr
p.Asn397Asp
75 kDa
SMPD1
45 kDa
B-actin
Empty Vector
SMPD1 WT
p.Ala44Pro
p.Pro282Thr
p.Asn337Ser
p.Tyr539His
p.Asp565Asn
p.Pro226Gln
75 kDa
SMPD1
45 kDa
B-actin

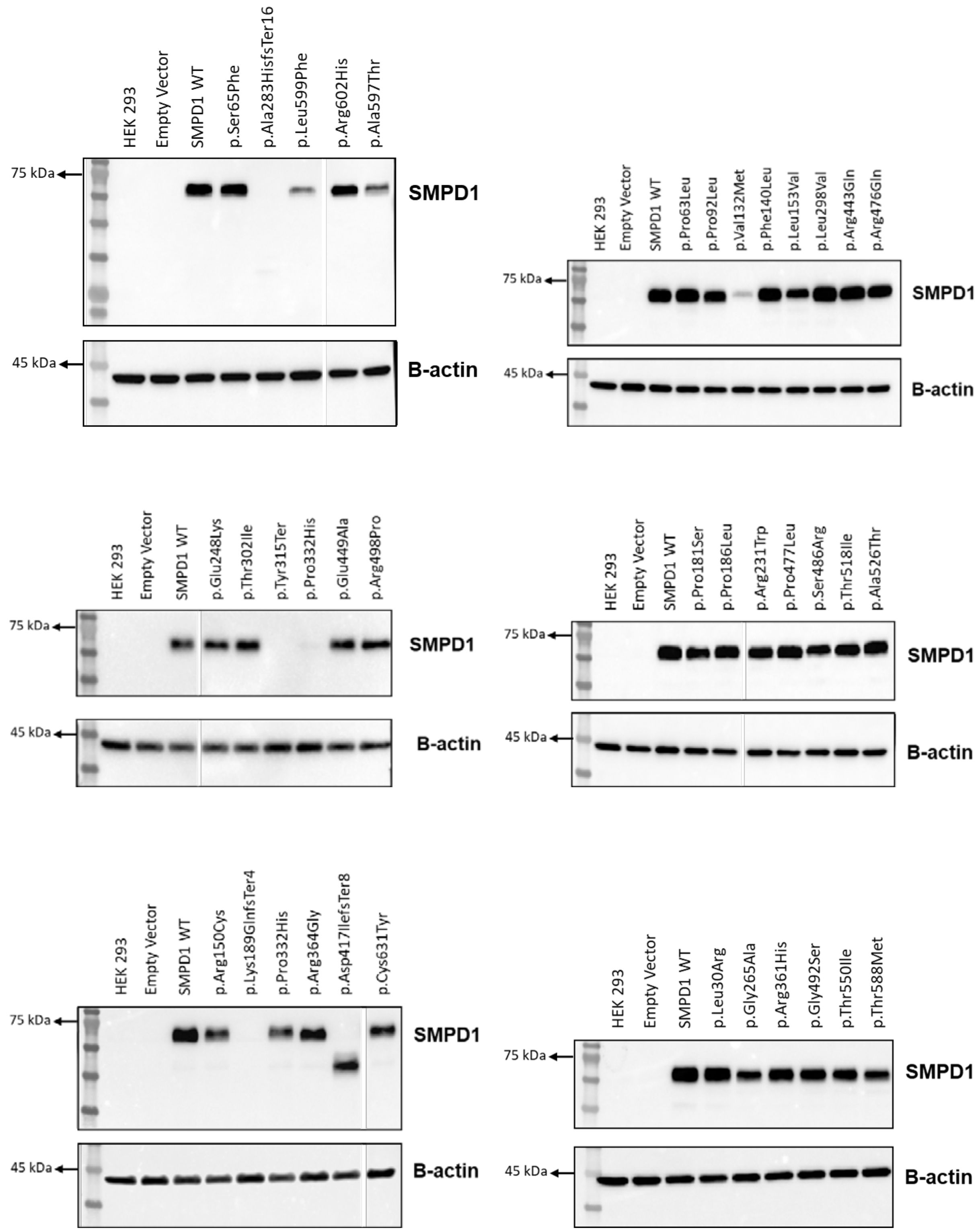
HEK 293
Empty Vector
SMPD1 WT
p.Ser65Phe
p.Ala283HisfsTer16
p.Leu599Phe
p.Arg602His
p.Ala597Thr
75 kDa
SMPD1
45 kDa
B-actin
HEK 293
Empty Vector
SMPD1 WT
p.Pro63Leu
p.Pro92Leu
p.Val132Met
p.Phe140Leu
p.Leu153Val
p.Leu298Val
p.Arg443Gln
p.Arg476Gln
75 kDa
SMPD1
45 kDa
B-actin
HEK 293
Empty Vector
SMPD1 WT
p.Glu248Lys
p.Thr302Ile
p.Tyr315Ter
p.Pro332His
p.Glu449Ala
p.Arg498Pro
75 kDa
SMPD1
45 kDa
B-actin
HEK 293
Empty Vector
SMPD1 WT
p.Pro181Ser
p.Pro186Leu
p.Arg231Trp
p.Pro477Leu
p.Ser486Arg
p.Thr518Ile
p.Ala526Thr
75 kDa
SMPD1
45 kDa
B-actin
HEK 293
Empty Vector
SMPD1 WT
p.Arg150Cys
p.Lys189GlnfsTer4
p.Pro332His
p.Arg364Gly
p.Asp417IlefsTer8
p.Cys631Tyr
75 kDa
SMPD1
45 kDa
B-actin
HEK 293
Empty Vector
SMPD1 WT
p.Leu30Arg
p.Gly265Ala
p.Arg361His
p.Gly492Ser
p.Thr550Ile
p.Thr588Met
75 kDa
SMPD1
45 kDa
B-actin

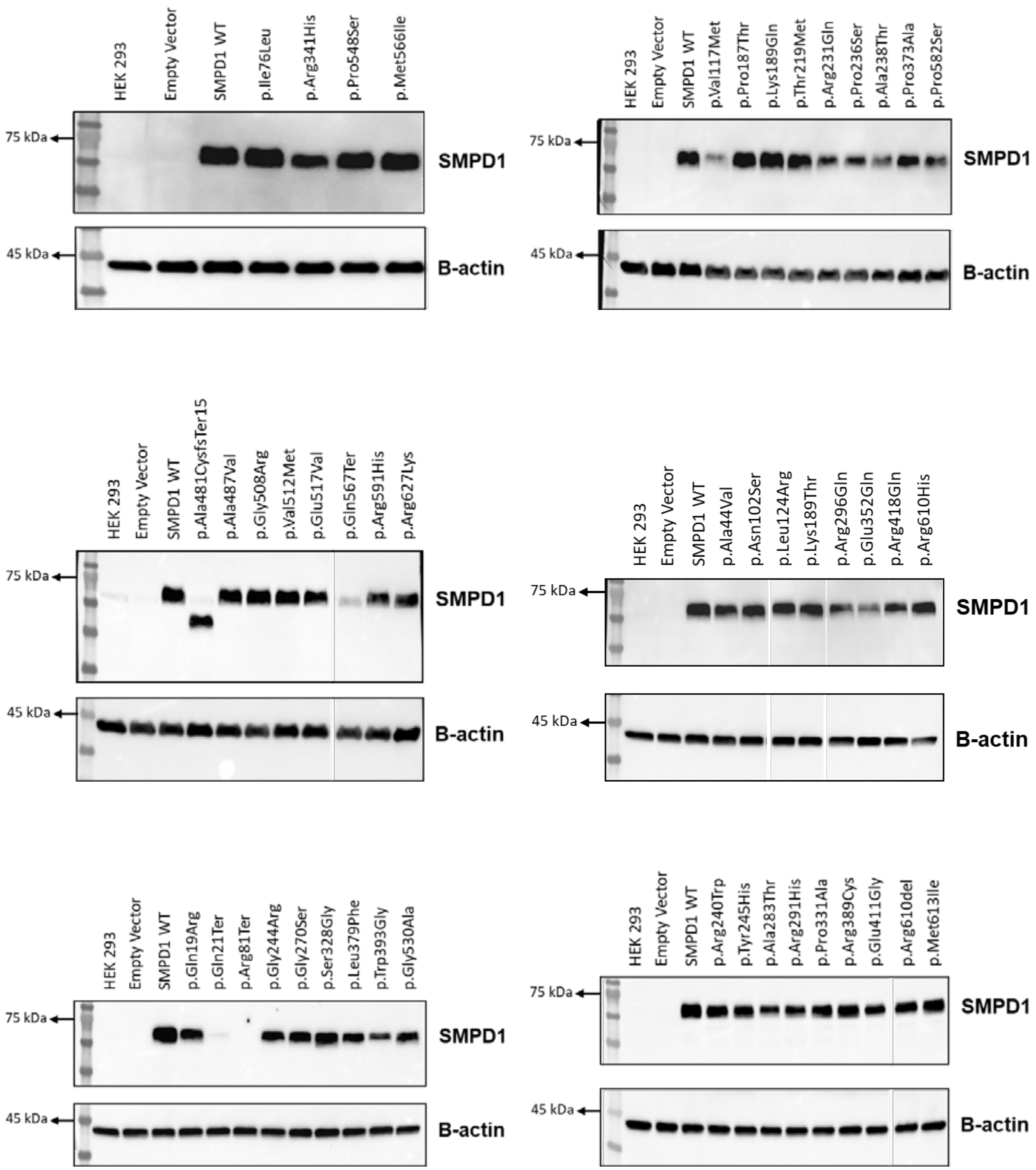
HEK 293
Empty Vector
SMPD1 WT
p.Ile76Leu
p.Arg341His
p.Pro548Ser
p.Met566Ile
75 kDa
SMPD1
45 kDa
B-actin
HEK 293
Empty Vector
SMPD1 WT
p.Val117Met
p.Pro187Thr
p.Lys189Gln
p.Thr219Met
p.Arg231Gln
p.Pro236Ser
p.Ala238Thr
p.Pro373Ala
p.Pro582Ser
75 kDa
SMPD1
45 kDa
B-actin
HEK 293
Empty Vector
SMPD1 WT
p.Ala481CysfsTer15
p.Ala487Val
p.Gly508Arg
p.Val512Met
p.Glu517Val
p.Gln567Ter
p.Arg591His
p.Arg627Lys
75 kDa
SMPD1
45 kDa
B-actin
HEK 293
Empty Vector
SMPD1 WT
p.Ala44Val
p.Asn102Ser
p.Leu124Arg
p.Lys189Thr
p.Arg296Gln
p.Glu352Gln
p.Arg418Gln
p.Arg610His
75 kDa
SMPD1
45 kDa
B-actin
HEK 293
Empty Vector
SMPD1 WT
p.Gln19Arg
p.Gln21Ter
p.Arg81Ter
p.Gly244Arg
p.Gly270Ser
p.Ser328Gly
p.Leu379Phe
p.Trp393Gly
p.Gly530Ala
75 kDa
SMPD1
45 kDa
B-actin
HEK 293
Empty Vector
SMPD1 WT
p.Arg240Trp
p.Tyr245His
p.Ala283Thr
p.Arg291His
p.Pro331Ala
p.Arg389Cys
p.Glu411Gly
p.Arg610del
p.Met613Ile
75 kDa
SMPD1
45 kDa
B-actin

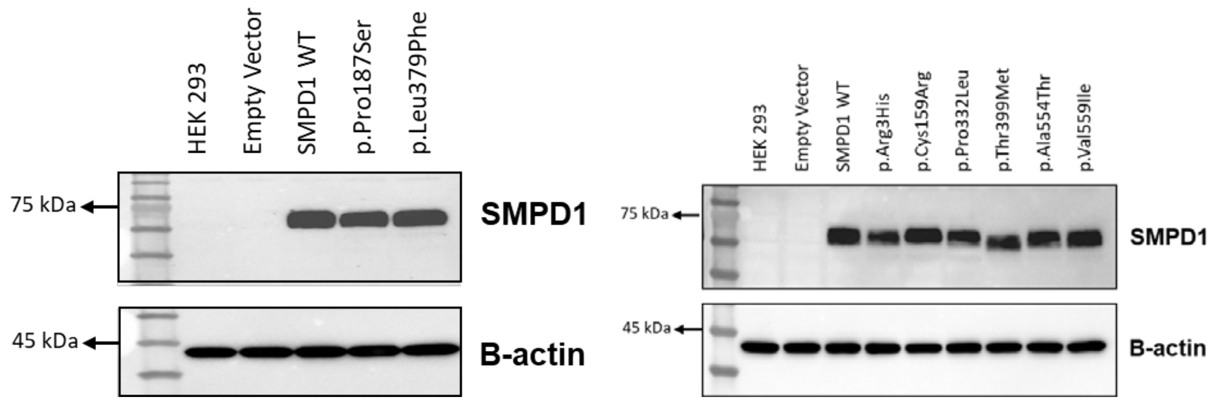


**Figure S9: Western blots of SMPD1 protein expression for all 125 assayed variants.**

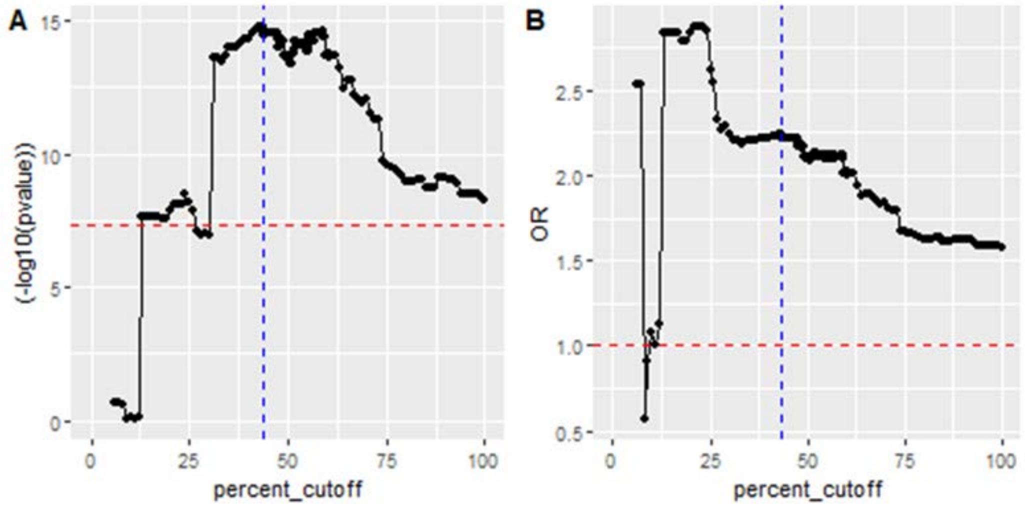


**Figure S10: Iterative linear discriminant analysis of *SMDP1* variants defined as functionally deficient at various ASM enzymatic activity thresholds.** The activity thresholds range from 0% to 100% relative to wild-type SMPD1 and are expressed as a continuous variable on the horizontal axis. Analysis was undertaken for all 9,883 persons with Parkinson's disease and 11,154 unaffected individuals described in this study.

The vertical axis for Panel A depicts the statistical significance ($-\log_{10}P$) of the association between burden of rare variants defined as functionally deficient and risk of Parkinson's disease. The dashed horizontal red line indicates $P = 5 \times 10^{-8}$.

The vertical axis for Panel B depicts the Odds Ratio (OR) of the association between burden of rare variants defined as functionally deficient and risk of Parkinson's disease. The dashed horizontal red line indicates OR = 1.

In both panels, the dashed vertical blue line indicates the ASM enzymatic activity level threshold of ≤ 43.58% in which the most significant difference in variant burden between PD cases and controls was observed ($P = 1.25 \times 10^{-15}$, OR = 2.24, 95% CI = 1.83 - 2.76).

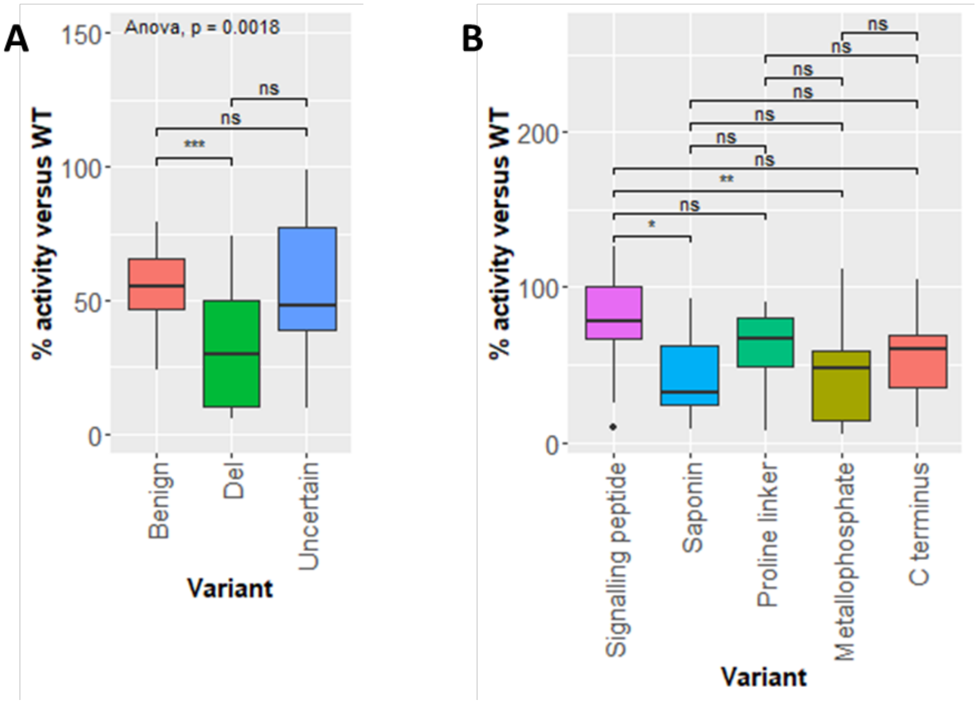


**Figure S11: Distribution of ASM enzymatic activity levels of rare *SMPD1* variants in relation to its (A) effects on tertiary structure integrity and (B) localization to protein domains.** Analysis for both panels are based on protein crystallography of ASM tertiary structure 5i81 [1].

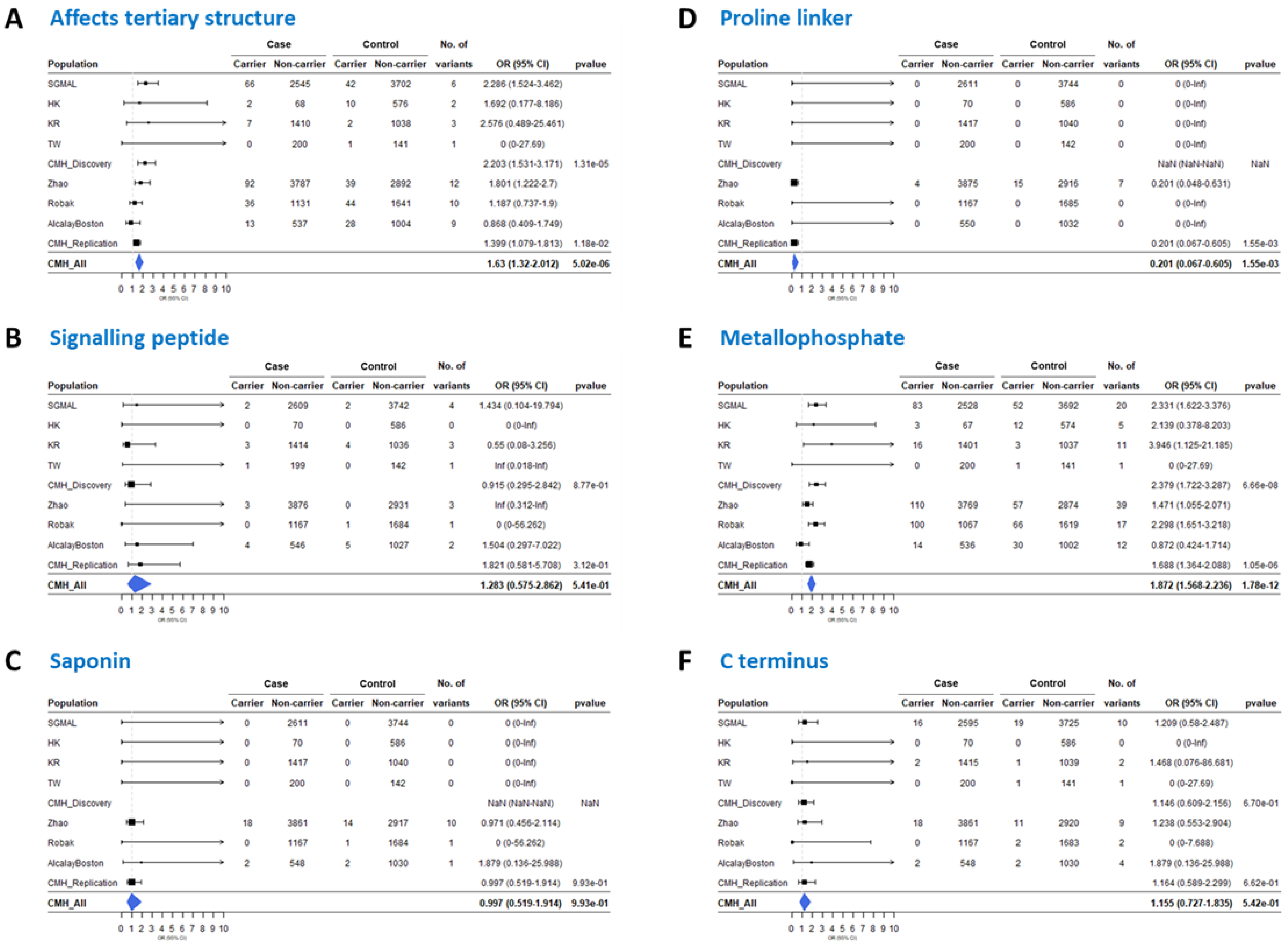
A Affects tertiary structure
Case Control No. of
Population Carrier Non-carrier Carrier Non-carrier variants OR (95% CI) pvalue
SGMAL 66 2545 42 3702 6 2.286 (1.524-3.462)
HK 2 68 10 576 2 1.692 (0.177-8.186)
KR 7 1410 2 1038 3 2.576 (0.489-25.461)
TW 0 200 1 141 1 0 (0-27.69)
CMH_Discovery 2.203 (1.531-3.171) 1.31e-05
Zhao 92 3787 39 2892 12 1.801 (1.222-2.7)
Robak 36 1131 44 1641 10 1.187 (0.737-1.9)
AlcalayBoston 13 537 28 1004 9 0.868 (0.409-1.749)
CMH_Replication 1.399 (1.079-1.813) 1.18e-02
CMH_All 1.63 (1.32-2.012) 5.02e-06
0 1 2 3 4 5 6 7 8 9 10
B Signalling peptide
Case Control No. of
Population Carrier Non-carrier Carrier Non-carrier variants OR (95% CI) pvalue
SGMAL 2 2609 2 3742 4 1.434 (0.104-19.794)
HK 0 70 0 586 0 0 (0-Inf)
KR 3 1414 4 1036 3 0.55 (0.08-3.256)
TW 1 199 0 142 1 Inf (0.018-Inf)
CMH_Discovery 0.915 (0.295-2.842) 8.77e-01
Zhao 3 3876 0 2931 3 Inf (0.312-Inf)
Robak 0 1167 1 1684 1 0 (0-56.262)
AlcalayBoston 4 546 5 1027 2 1.504 (0.297-7.022)
CMH_Replication 1.821 (0.581-5.708) 3.12e-01
CMH_All 1.283 (0.575-2.862) 5.41e-01
0 1 2 3 4 5 6 7 8 9 10
C Saponin
Case Control No. of
Population Carrier Non-carrier Carrier Non-carrier variants OR (95% CI) pvalue
SGMAL 0 2611 0 3744 0 0 (0-Inf)
HK 0 70 0 586 0 0 (0-Inf)
KR 0 1417 0 1040 0 0 (0-Inf)
TW 0 200 0 142 0 0 (0-Inf)
CMH_Discovery NaN (NaN-NaN) NaN
Zhao 18 3861 14 2917 10 0.971 (0.456-2.114)
Robak 0 1167 1 1684 1 0 (0-56.262)
AlcalayBoston 2 548 2 1030 1 1.879 (0.136-25.988)
CMH_Replication 0.997 (0.519-1.914) 9.93e-01
CMH_All 0.997 (0.519-1.914) 9.93e-01
0 1 2 3 4 5 6 7 8 9 10
D Proline linker
Case Control No. of
Population Carrier Non-carrier Carrier Non-carrier variants OR (95% CI) pvalue
SGMAL 0 2611 0 3744 0 0 (0-Inf)
HK 0 70 0 586 0 0 (0-Inf)
KR 0 1417 0 1040 0 0 (0-Inf)
TW 0 200 0 142 0 0 (0-Inf)
CMH_Discovery NaN (NaN-NaN) NaN
Zhao 4 3875 15 2916 7 0.201 (0.048-0.631)
Robak 0 1167 0 1685 0 0 (0-Inf)
AlcalayBoston 0 550 0 1032 0 0 (0-Inf)
CMH_Replication 0.201 (0.067-0.605) 1.55e-03
CMH_All 0.201 (0.067-0.605) 1.55e-03
0 1 2 3 4 5 6 7 8 9 10
E Metallophosphate
Case Control No. of
Population Carrier Non-carrier Carrier Non-carrier variants OR (95% CI) pvalue
SGMAL 83 2528 52 3692 20 2.331 (1.622-3.376)
HK 3 67 12 574 5 2.139 (0.378-8.203)
KR 16 1401 3 1037 11 3.946 (1.125-21.185)
TW 0 200 1 141 1 0 (0-27.69)
CMH_Discovery 2.379 (1.722-3.287) 6.66e-08
Zhao 110 3769 57 2874 39 1.471 (1.055-2.071)
Robak 100 1067 66 1619 17 2.298 (1.651-3.218)
AlcalayBoston 14 536 30 1002 12 0.872 (0.424-1.714)
CMH_Replication 1.688 (1.364-2.088) 1.05e-06
CMH_All 1.872 (1.568-2.236) 1.78e-12
0 1 2 3 4 5 6 7 8 9 10
F C terminus
Case Control No. of
Population Carrier Non-carrier Carrier Non-carrier variants OR (95% CI) pvalue
SGMAL 16 2595 19 3725 10 1.209 (0.58-2.487)
HK 0 70 0 586 0 0 (0-Inf)
KR 2 1415 1 1039 2 1.468 (0.076-86.681)
TW 0 200 1 141 1 0 (0-27.69)
CMH_Discovery 1.146 (0.609-2.156) 6.70e-01
Zhao 18 3861 11 2920 9 1.238 (0.553-2.904)
Robak 0 1167 2 1683 2 0 (0-7.688)
AlcalayBoston 2 548 2 1030 4 1.879 (0.136-25.988)
CMH_Replication 1.164 (0.589-2.299) 6.62e-01
CMH_All 1.155 (0.727-1.835) 5.42e-01
0 1 2 3 4 5 6 7 8 9 10

**Figure S12: Forest plot of rare *SMPD1* variants from Discovery and Replication datasets that (A) affect ASM tertiary structure, (B) localized to the signaling peptide, (C) localized to the signaling peptide, (D) localized to the proline-rich linker, (E) localized to the catalytic metallophosphate domain, and (F) localized to the C terminus.** Effect of variants on ASM tertiary structure and localisation based on protein crystallography of ASM tertiary structure 5i81 [1].

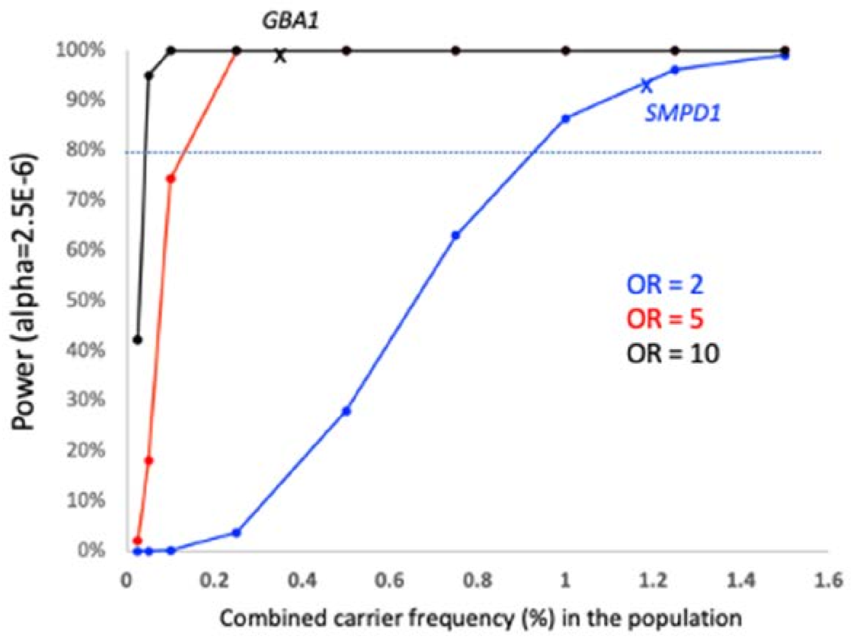


**Figure S13: Statistical power to identify PD-associated genes at exome-wide significance.**

Statistical power[6] to identify PD-associated genes at exome-wide significance (alpha=$2.5\times10^{-6}$) is dependent on sample size, the combined carrier frequency in the population and effect size (odds ratio) of the variants in each gene.

Based on our current discovery cohort size of 4,298 PD cases and 5,512 controls, we had >90% power to detect genes like *GBA1* with large effect sizes (OR>5) and at combined carrier frequencies ~0.5% in the population, and also genes like *SMPD1* with smaller effect size (OR ~2) but at higher combined carrier frequencies of 1-1.5% in the population (contributed in part by the *SMPD1* p.Pro332Arg variant). Genes with similar variant effect sizes as *SMPD1* may be missed if they are present at lower combined carrier frequencies of 0.9% or less. Similarly, genes with variant effect sizes comparable with *GBA1* will be missed if they are present at combined carrier frequencies of 0.1% or less.
